# PNEUMOCOCCAL SEROTYPE DISTRIBUTION AND COVERAGE OF EXISTING AND PIPELINE PNEUMOCOCCAL VACCINES

**DOI:** 10.1101/2024.12.12.24318944

**Authors:** Laura M King, Kristin L Andrejko, Miwako Kobayashi, Wei Xing, Adam L Cohen, Wesley H Self, J Jackson Resser, Cynthia G Whitney, Adrienne Baughman, Mai Kio, Carlos G Grijalva, Jessica Traenkner, Nadine Rouphael, Joseph A Lewnard

## Abstract

**Background:** *Streptococcus pneumoniae* (pneumococcus) causes invasive pneumococcal disease (IPD) and non-invasive acute respiratory infections (ARIs). Three pneumococcal conjugate vaccines (PCVs) are recommended in the United States with additional products in clinical trials. We aimed to estimate 1) proportions of IPD cases and pneumococcal ARIs caused by serotypes targeted by existing and pipeline PCVs and 2) annual U.S. pneumococcal burdens potentially preventable by PCVs.

**Methods:** We estimated serotype distribution and proportions of non-invasive pneumococcal ARIs (AOM [children only], sinusitis, non-bacteremic pneumonia) and IPD attributable to serotypes targeted by each PCV using Markov chain Monte Carlo approaches incorporating data from studies of serotype distribution in ARIs and Active Bacterial Core Surveillance (ABCs) data. We then estimated annual numbers of outpatient-managed pneumococcal ARIs, non-bacteremic pneumococcal pneumonia hospitalizations, and IPD cases potentially preventable by PCVs in the United States by multiplying pneumococcal disease incidence rates by PCV-targeted proportions of disease and vaccine effectiveness estimates.

**Results:** In children, PCV15, PCV20, PCV24, PCV25, and PCV31 serotypes account for 16% (95% confidence interval: 15–17%), 31% (30–32%), 34% (32–35%), 43% (42–44%), and 68% (67–69%) of pneumococcal acute otitis media cases, respectively. In adults, PCV15, PCV20, PCV21, PCV24, PCV25, and PCV31 serotypes account for 43% (38–47%), 52% (47–57%), 69% (64–73%), 65% (61–70%), 62% (57–67%), and 87% (83–90%) of pneumococcal non-bacteremic pneumonia cases. For IPD, 42–85% of pediatric and 42–94% of adult cases were due to PCV-targeted serotypes. PCV-preventable burdens encompassed 270 thousand–3.3 million outpatient-managed ARIs, 2–17 thousand non-bacteremic pneumonia hospitalizations, and 3–14 thousand IPD cases in the United States annually.

**Conclusions:** Across pneumococcal conditions, coverage and preventable burdens were lowest for PCV15 and highest for PCV31, with PCV21 also targeting sizeable burdens of adult disease. Serotype distribution across syndromes may inform vaccine formulations and policy.

## INTRODUCTION

*Streptococcus pneumoniae* (pneumococcus) causes invasive pneumococcal disease (IPD; bacteremia, meningitis, bacteremic pneumonia) and non-invasive acute respiratory infections (ARIs), including acute otitis media (AOM), sinusitis, and non-bacteremic pneumonia. Over 100 pneumococcal serotypes have been identified via polysaccharide capsule antigens,^1^ with a minority important in human disease. IPD serotype distribution is well-documented.^2,3^ However, little is known about serotype distribution in pneumococcal ARIs despite large burdens of ARIs in U.S. children and adults.^4,5^

Pneumococcal conjugate vaccines (PCVs) reduce vaccine-serotype colonization and disease.^6–8^ In the United States, 7-valent PCV (PCV7) was introduced in 2000 followed by 13-valent PCV (PCV13) in 2010. Currently, the Advisory Committee on Immunization Practices (ACIP) recommends 15-and 20-valent PCVs (PCV15, PCV20) for infants^9,10^ and 21-valent PCV (PCV21) alone, PCV20 alone, or PCV15 plus 23-valent pneumococcal polysaccharide vaccine (PPSV23) for PCV-naïve adults aged ≥50 years and those 19–49 years with certain medical conditions.^11,12^ PCV-induced pressure on vaccine-targeted serotypes results in increased non-vaccine serotype circulation,^13–15^ necessitating periodic PCV reformulation (serotype addition and removal). At present, four pipeline pneumococcal vaccines are undergoing clinical trials: 24-valent PCV (PCV24), 24-valent Pn-MAPS24v (employs Multiple Antigen Presenting System platform; henceforth grouped with PCV24), 25-valent PCV (PCV25), and 31-valent PCV (PCV31) (**Figure 1**). Vaccine formulations are largely informed by serotypes in IPD; it is unknown to what extent current and pipeline PCVs may contribute to reductions in ARIs.

**Figure 1.**
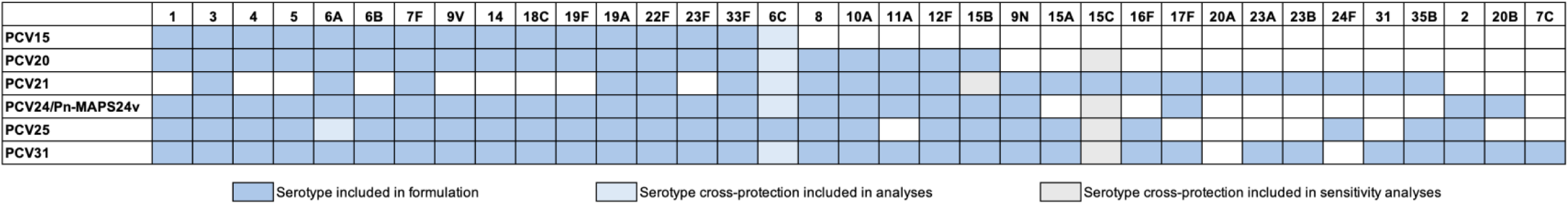
Pneumococcal serotypes targeted by pneumococcal conjugate vaccine products. The 24-valent Pn-MAPS24v uses a Multiple Antigen Presenting System platform rather than the traditional conjugate protein platform. PCV24 and Pn-MAPS24v target the same serotypes and so are considered together.

We aimed to estimate serotype distribution and corresponding PCV-targeted proportions of IPD and non-invasive pneumococcal ARIs in U.S. children and adults using Markov chain Monte Carlo (MCMC) approaches incorporating data from studies of serotype distribution in these syndromes. We used these results to model disease burdens potentially preventable by existing and pipeline PCVs in the United States.

## METHODS

We estimated serotype distribution and proportions of non-invasive pneumococcal ARIs (AOM [children only], sinusitis, non-bacteremic pneumonia) and IPD attributable to serotypes targeted by each PCV. We then estimated potential vaccine-preventable burdens by multiplying pneumococcal disease incidence rates by PCV-targeted proportions of disease and vaccine effectiveness (VE) estimates. **Table S1** summarizes data inputs.

### Serotype distribution in pediatric ARIs

We conducted a literature review of studies from the PubMed database (**Table S2**) with data on pneumococcal serotypes in nasopharyngeal samples from children with ARIs in high-income countries after PCV13 implementation. We excluded studies not in English, from countries using multiple PCVs, and that aggregated data from children with and without ARI.

We conducted meta-analyses of identified studies (**Table S3**) to estimate serotype distributions among pneumococcal isolates sampled from children with 1) AOM, and 2) any ARI. No post-PCV13 studies evaluated sinusitis etiology and only one evaluated pneumonia etiology.^16^ Thus, parameterization of pneumococcal sinusitis and non-bacteremic pneumonia serotype distributions relied on data from one pneumonia and AOM studies. We considered all serotypes except 15D,^17^ only identified in IPD, resulting in 100 serotype categories, including a non-typeable category.

For 1) AOM and 2) other ARIs, we estimated serotype-specific proportions using a Markov chain Monte Carlo (MCMC) approach drawing on serotype-specific frequencies and sample sizes in each study. We defined **p** as a vector of serotype-specific prevalences (*p*_*i*_ for *i* ∈ {1, … ,100}) among all pneumococcal isolates, such that 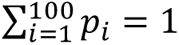. For each study *j*, we defined the vector of serotype-specific frequencies **x**_*j*_ = {*x*_1,*j*_, *x*_2,*j*_, … , *x*_100,*j*_} as a multinomial draw parameterized by serotype distributions across studies and the sample size *n*_*j*_ = ∑_*i*_ *x*_*i*,*j*_,

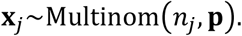

Where studies aggregated data across serotypes/serogroups or did not report all serotypes, we summed corresponding *p*_*i*,*j*_ values to align with the categories represented by the reported **x**_*j*_ data. We applied the correction

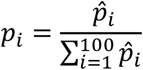

to the **p̑** proposal distribution to ensure that serotype prevalences summed to 1. We conducted 5,000,000 draws with 100,000 draw burn-ins.

### Serotype distribution in IPD

We used serotype-specific counts from 2015–2019 Active Bacterial Core surveillance (ABCs) data^18^ to parameterize IPD serotype distributions. We stratified serotype-specific IPD data by age category (0– 17, 18–49, 50–64, and ≥65 years) and sampled serotype distribution vectors via MCMC, parameterized according to multinomial serotype frequencies. For IPD, we included 15D, thus considering 101 serotypes.

### Serotype distribution in adult non-bacteremic pneumonia

We used serotype-specific counts from the Pneumococcal pNeumonia Epidemiology, Urine serotyping, and Mental Outcomes (PNEUMO) study^19^ supplemented with ABCs data to parameterize serotype distributions in adult non-bacteremic pneumococcal pneumonia. The PNEUMO study used a novel serotype-specific urinary antigen detection (SSUAD) assay to identify 30 pneumococcal serotypes: 1, 3, 4, 5, 6A/C, 6B, 7F, 8, 9N, 9V, 10A, 11A, 12F, 14, 15A, 15C, 16F, 17F, 18C, 19A, 19F, 20A, 22F, 23A, 23B, 23F, 24F, 31, 33F, and 35B. In the study population, 20% of patients (69/352) with pneumococcal pneumonia did not test positive for SSUAD-identified serotypes.

For the 30 SSUAD serotypes, we used MCMC to estimate a serotype-specific prevalence vector **π** subset to SSUAD serotypes from data among the 283 patients with SSUAD-serotype pneumonia. We combined 6A/C due to assay cross-reactivity. For non-SSUAD serotypes, we assumed their distribution resembled that in adult IPD due to non-SSUAD serotypes:

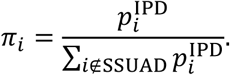

We multiplied resulting serotype-specific prevalence estimates by proportions of patients experiencing non-bacteremic pneumococcal pneumonia associated with SSUAD and non-SSUAD serotypes:

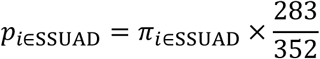

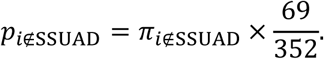

### PCV-targeted pneumococcal disease

By condition, we defined coverage for each PCV as the sum of estimated prevalences of targeted serotypes. In primary analyses, we only considered PCV21 in adults due to its adult-only indication.^20^ We conducted sensitivity analyses for PCV21-preventable burdens of disease among children at increased risk of pneumococcal disease given current clinical trials.^21^ We considered 6A/C together, consistent with previous studies,^22^ due to demonstrated cross-protection^23,24^ and SSUAD cross-reactivity.^19,25^ We considered 15B/C separately in primary analyses and together in sensitivity analyses. Although vaccination with PCV20 led to detection of anti-15C antibody in adults^26^ and children^27^, clinically-relevant cross-protection is unknown.

### Preventable disease burdens

We estimated direct preventable disease burdens as the products of pneumococcal disease incidence, VE estimates, and PCV serotype coverage by condition and age group.

We used multiple sources to estimate the incidence of pneumococcal ARIs and IPD. When possible, we used 2019 data to capture the most recent data without impact from COVID-19 pandemic-associated shifts in disease transmission and healthcare-seeking behavior. For IPD, we obtained incidence estimates from ABCs data.^28^ For pneumococcal ARIs, we estimated burdens by multiplying all-cause AOM, pneumonia, and sinusitis incidence rates by published estimates of pneumococcal-attributable proportions of cases (**Table S4**). For non-bacteremic pneumonia, we generated separate estimates by inpatient and outpatient settings. National counts of 2019 inpatient all-cause pneumonia were estimated using National Inpatient Sample (NIS)^29^ data. To obtain non-bacteremic pneumococcal pneumonia hospitalizations, we subtracted estimated bacteremic pneumococcal pneumonia cases from total pneumococcal hospitalization estimates. Bacteremic pneumococcal pneumonia cases were estimated by multiplying national IPD case counts by the proportion that are bacteremic pneumonia (72.1%).^28^ We assumed outpatient-managed pneumonia was non-bacteremic. We estimated AOM and sinusitis burdens using outpatient visit incidence. Estimates of outpatient-managed AOM, pneumonia, and sinusitis were derived from 2016 and 2019 National Ambulatory and National Hospital Ambulatory Medical Care Surveys (NAMCS/NHAMCS) and 2016–2019 Meritage MarketScan Commercial and Medicaid databases following previously-described methods.^30,31^ We used the ratio of visit incidence in NAMCS/NHAMCS and MarketScan among adults aged 50–64 years to impute MarketScan incidence rates in adults ≥65 years. Only 2016 and 2019 NAMCS/NHAMCS data were used due to data limitations in 2017-18 datasets.^32^ For outpatient-managed pneumonia, we stratified pediatric age groups as <2 and 2–17 years to ensure adequate sample size for NAMCS/NHAMCS projection validity.^32^ We propagated uncertainty by fitting age-and condition-specific burden estimates to Gamma distributions. We multiplied incidence rates by 2019 bridged-race census estimates^33^ to obtain national counts.

For age groups (5–17, 18–49 years) for which PCVs are only recommended for individuals at increased risk of pneumococcal disease,^9–11,34^ we estimated burdens among 1) all individuals, and 2) those with risk conditions, using published estimates (**Table S5**).

PCV licensure is based on immune response non-inferiority; clinical protection estimates are not yet available for PCVs in this study. Age-specific vaccine-serotype VE estimates were extracted from studies of PCV7 and PCV13 for all conditions except pediatric non-bacteremic pneumonia (**Table S6**). Although protective against non-bacteremic pneumonia in children,^35^ PCV VE against vaccine-serotype disease is uncertain.^36,37^ Consistent with prior work,^38^ we obtained VE against vaccine-serotype non-bacteremic pneumonia in children by multiplying VE against vaccine-serotype pediatric IPD by the ratio of vaccine-serotype VE estimates in adults for non-bacteremic pneumonia and IPD (45%:75%).^39^ We propagated uncertainty by fitting Beta distributions to published estimates.

For preventable burden estimates, we excluded PCV13 serotypes from PCV-targeted proportions of disease assuming that residual PCV13-serotype disease was not further preventable by higher-valency PCVs. We did not consider indirect effects.

This activity was reviewed by CDC, deemed non-human-subjects, and conducted consistent with applicable federal law and CDC policy. See e.g., 45 C.F.R. part 46.102(l)(2), 21 C.F.R. part 56; 42 U.S.C. §241(d); 5 U.S.C. §552a; 44 U.S.C. §3501 et seq.

## RESULTS

### Serotype distribution

Estimated via meta-analysis, the most prevalent serotypes in pediatric pneumococcal ARIs were 15C, 23B, 11A, 15A, 35B, and 23A, together accounting for >50% of isolates (**Figure 2**; **Table S7**). Serotypes 3, 22F, 20B, 19A, 35B, 9N, 19F and 23A were the most frequent causes of adult non-bacteremic pneumococcal pneumonia. Serotypes 6A/C were common in adults aged ≥65 years but infrequently identified among adults <65 years. Serotype 3 accounted for 1.4% (95% confidence interval 1.1–1.7%) of pneumococci in pediatric ARIs and 11.6% (8.7–15.0%) in adult non-bacteremic pneumonia.

**Figure 2.**
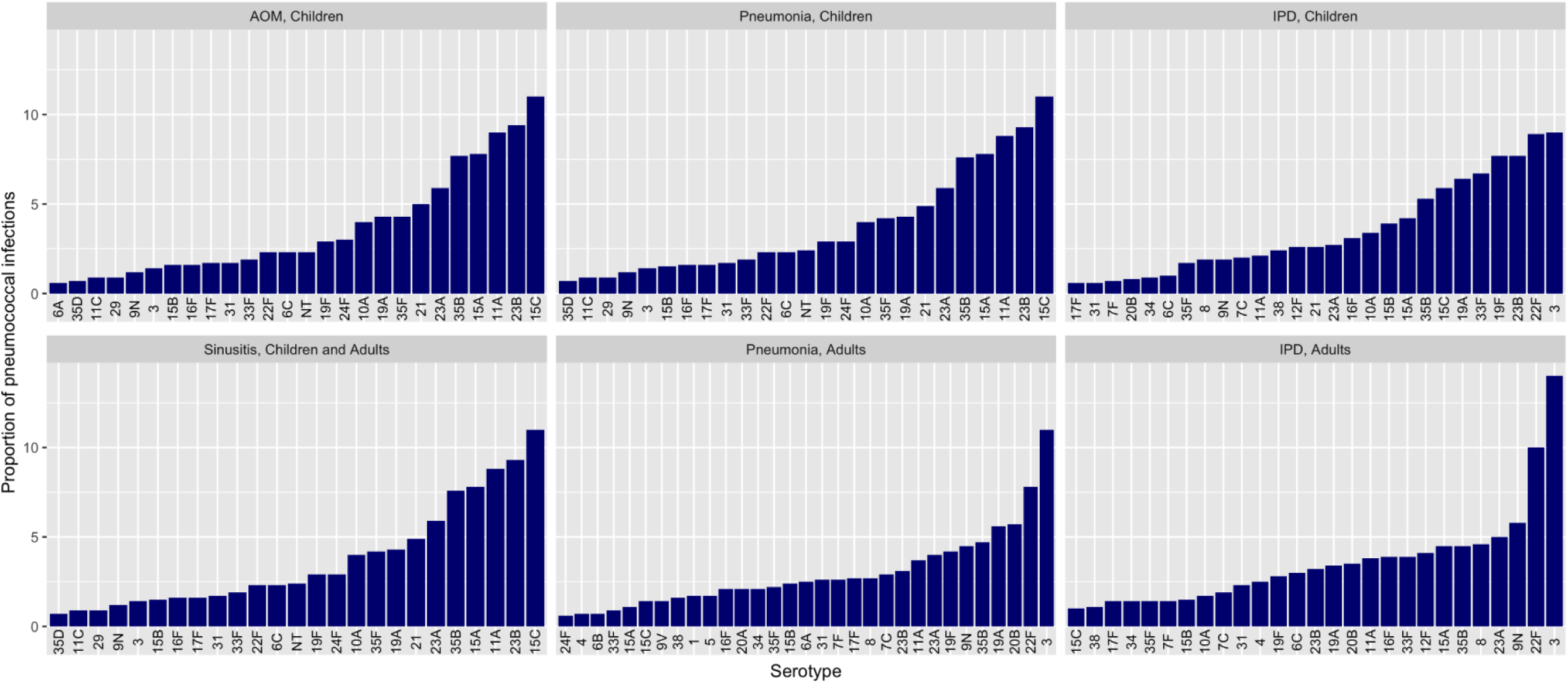
Serotype-specific proportions of pneumococcal infections by condition and age group. Serotypes accounting for <0.05% of pneumococcal isolates within a condition and age group not displayed. All serotype distribution estimates detailed in **Table S7**.

Serotype 3 was the most prevalent serotype in IPD, followed by 22F (**Figure 2**; **Table S7)**. In pediatric IPD, the 10 most prevalent serotypes (3, 22F, 19F, 23B, 33F, 19A, 15C, 35B, 15A, 15B) accounted for almost two-thirds of all cases. In adult IPD, 9N was the third most common serotype, accounting for 5.8% (5.4–6.2%) of cases.

### PCV pneumococcal disease coverage

In children, PCV15, PCV20, PCV24, PCV25 and PCV31 serotypes accounted for 16.1% (15.2–17.0%), 30.7% (29.6–31.8%), 33.6% (32.4–34.8%), 43.0% (41.8–44.2%), and 68.0% (66.8–69.2%) of pneumococcal AOM cases, respectively (**Table 1**). Serotype distribution in pneumococcal sinusitis and non-bacteremic pneumonia and were similar. For pediatric IPD, PCV15 serotypes accounted for 41.7% (38.9–44.7%) of cases, PCV20, PCV24, and PCV25 serotypes caused 56–68% of cases, and PCV31 serotypes accounted for 85.0% (82.6–87.1%) of cases. Additionally, 75.3% (72.6–77.8%) of pediatric IPD cases were due to serotypes in PCV21 (**Table S8**), currently in clinical trials for use in children with risk-based pneumococcal vaccine indications.

**Table 1.** Proportion of pneumococcal infections in children 0–17 years due to serotypes targeted by pneumococcal conjugate vaccine products^a,b,c^.

| Condition | Percent of pneumococcal isolates (95% CI) |  |  |  |  |
| --- | --- | --- | --- | --- | --- |
|  | PCV15 | PCV20 | PCV24<sup>d</sup> | PCV25 | PCV31 |
| AOM | 16.1 (15.2, 17.0) | 30.7 (29.6, 31.8) | 33.6 (32.4, 34.8) | 43.0 (41.8, 44.2) | 68.0 (66.8, 69.2) |
| Sinusitis, pneumonia<sup>e,f</sup> | 16.7 (15.9, 17.7) | 31.2 (30.1, 32.4) | 34.2 (33.0, 35.3) | 43.6 (42.4, 44.8) | 68.2 (67.1, 69.4) |
| IPD | 41.7 (38.9, 44.7) | 55.9 (52.9, 58.9) | 59.3 (56.4, 62.3) | 68.2 (65.3, 71.0) | 85.0 (82.6, 87.1) |

Abbreviations: CI – confidence interval; PCV – pneumococcal conjugate vaccine; AOM – acute otitis media; IPD – invasive pneumococcal disease.
<sup>a</sup> PCV15 includes serotypes 1, 3, 4, 5, 6A/C, 6B, 7F, 9V, 14, 18C, 19F, 19A, 22F, 23F, 33F. PCV20 includes serotypes 1, 3, 4, 5, 6A/C, 6B, 7F, 8, 9V, 10A, 11A, 12F, 14, 15B, 18C, 19F, 19A, 22F, 23F, 33F. PCV24 includes serotypes 1, 2, 3, 4, 5, 6A/C, 6B, 7F, 8, 9N, 9V, 10A, 11A, 12F, 14, 15B, 17F, 18C, 19F, 19A, 20B, 22F, 23F, 33F. PCV25 includes serotypes 1, 2, 3, 4, 5, 6B, 6A/C, 7F, 8, 9N, 9V, 10A, 12F, 14, 15A, 15B, 16F, 18C, 19F, 19A, 22F, 23F, 24F, 33F, 35B. PCV31 includes serotypes 1, 2, 3, 4, 5, 6A/C, 6B, 7F, 7C, 8, 9N, 9V, 10A, 11A, 12F, 14, 15A, 15B, 16F, 17F, 18C, 19F, 19A, 20B, 22F, 23F, 23A, 23B, 31, 33F, 35B.
<sup>b</sup> Assuming no 15B and 15C cross-protection.
<sup>c</sup> PCV21 not considered as it is intended only for use in adults. Sensitivity analyses considering PCV21 coverage for children with risk-based indications for PCVs presented in **Table S8**.
<sup>d</sup> Also includes Pn-MAPS24v.
<sup>e</sup> Non-bacteremic pneumonia. Bacteremic pneumonia included in IPD definition.
<sup>f</sup> Sinusitis and pneumonia serotype distribution and PCV coverage estimates derived from meta-analysis of serotype distribution in pediatric AOM and pneumonia.

In adults, PCV15 and PCV20 serotypes accounted for 42.6% (37.8–47.1%) and 52.1% (47.3–56.7%) of non-bacteremic pneumococcal pneumonia while PCV21, PCV24, and PCV25 serotypes accounted for 62–69% and PCV31 serotypes accounted for 86.7% (82.6–89.9%; **Table 2**). Proportions of cases targeted by PCV15 and PCV20 were higher among adults aged ≥65 years compared with those 18–49 years (44–56% versus 37–45%). In contrast, PCV21-targeted serotypes were more prevalent among younger adults than older adults. In adult IPD, PCV15, PCV20, PCV21, PCV24, PCV25, and PCV31 targeted 42.3% (41.4–43.1%), 58.1% (57.2–58.9%), 82.0% (81.3–82.7%), 68.8% (68.0–69.6%), 72.9% (72.2–73.7%), and 93.9% (93.5–94.3%) of cases. For all PCVs, except PCV21, IPD coverage was inversely related to age.

**Table 2.**
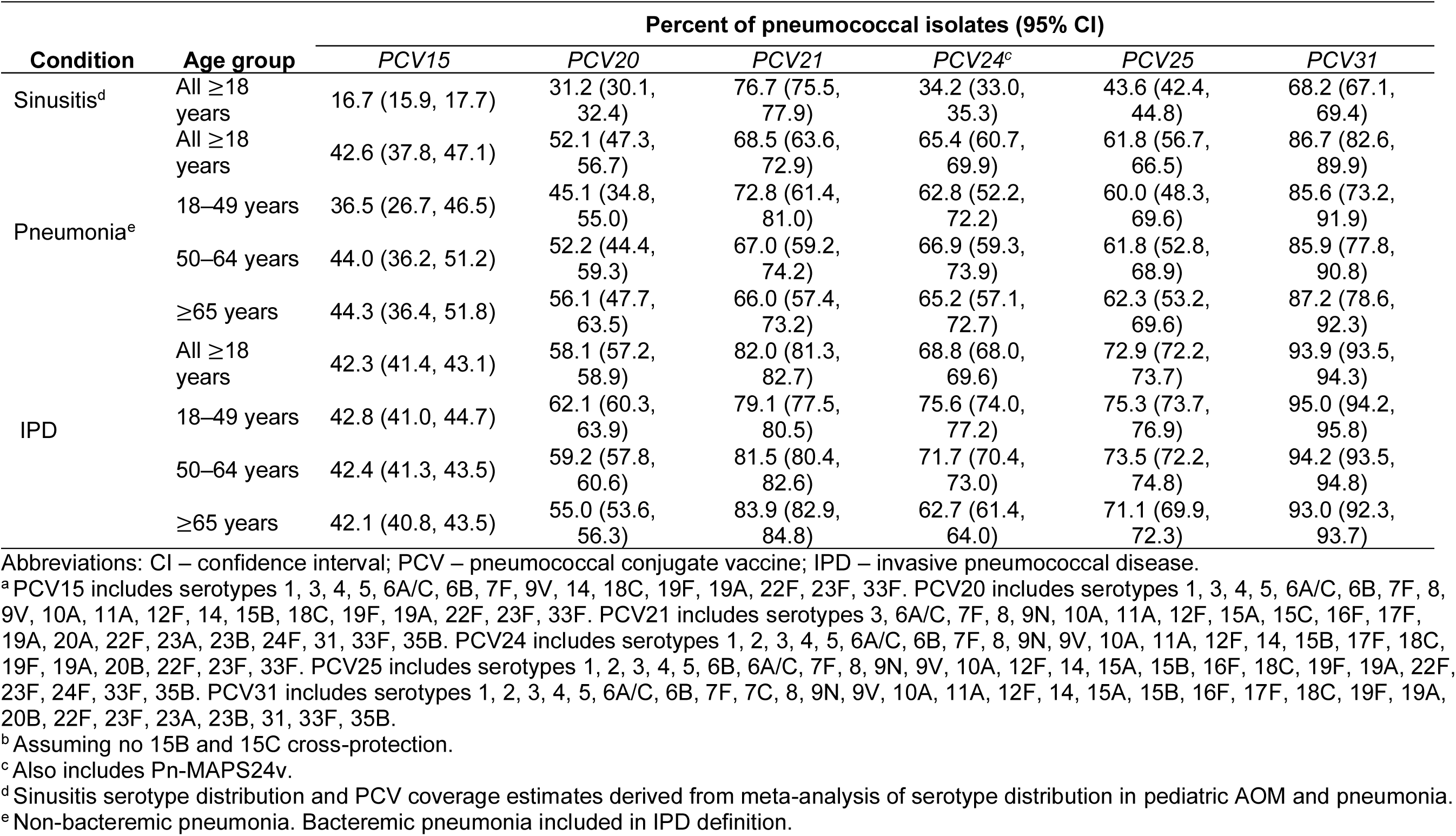
Proportion of pneumococcal infections in adults ≥18 years due to serotypes targeted by pneumococcal conjugate vaccine products^a,b^.

Pediatric coverage estimates increase markedly under the assumption that 15B-targeting vaccines (PCV20, PCV24, PCV25, PCV31) confer cross-protection against 15C (**Table S9**) while adult estimates increased slightly (**Table S10**). Serotype 15C accounts for 11.4% (10.5–12.2%) of pediatric pneumococcal ARIs and 5.9% (4.6–7.4%) of pediatric IPD, but <2% of adult pneumococcal pneumonia and approximately 1% of adult IPD (varying by age group; **Figure 2**, **Table S7**).

### Potentially preventable burdens

Accounting for all-cause disease burdens (**Tables S11–S13**), preventable (non-PCV13) serotype coverage (**Tables S14-S15**), and VE (**Table S6**), PCV-preventable disease burdens among U.S. children encompassed 58,189–784,825 outpatient-managed ARIs, 265–3,547 non-bacteremic pneumonia hospitalizations, and 235–888 IPD cases annually (**Table 3**, **Table S16**). The greatest potentially preventable burdens were from PCV31. When 15B/C cross-protection was considered, preventable burdens of pediatric pneumococcal ARIs increased notably while preventable pediatric IPD estimates increased slightly (**Table S17**). Limited to children with only risk-based PCV indications, PCV21 could prevent 203,136 (138,227–293,061) outpatient-managed ARIs, 499 (296–757) non-bacteremic pneumonia hospitalizations, and 130 (103–157) IPD cases (**Table S8**).

**Table 3.**
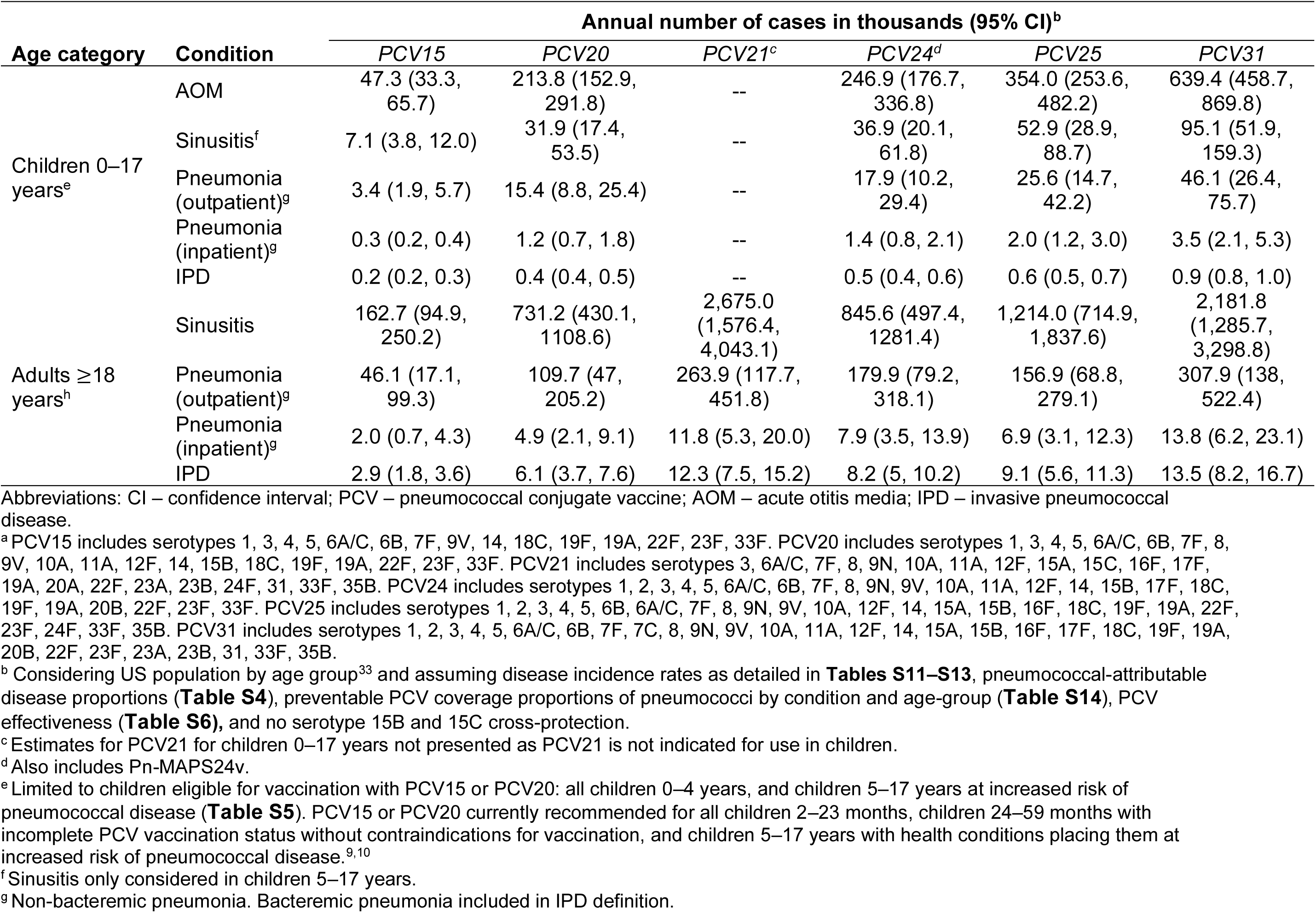
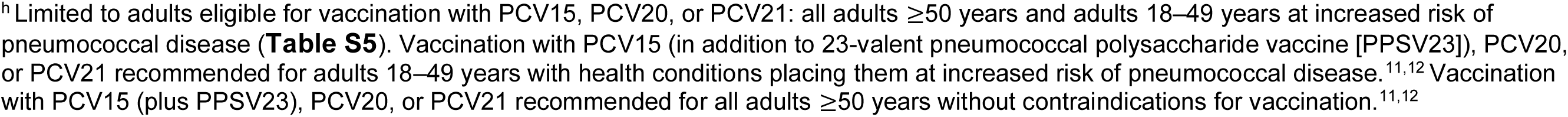
Estimated annual number of pneumococcal disease cases potentially preventable by pneumococcal conjugate vaccines^a^ in the United States.

| Age category | Condition | Annual number of cases in thousands (95% CI)<sup>b</sup> |  |  |  |  |  |
| --- | --- | --- | --- | --- | --- | --- | --- |
|  |  | PCV15 | PCV20 | PCV21<sup>c</sup> | PCV24<sup>d</sup> | PCV25 | PCV31 |
| Children 0–17 years<sup>e</sup> | AOM | 47.3 (33.3, 65.7) | 213.8 (152.9, 291.8) | -- | 246.9 (176.7, 336.8) | 354.0 (253.6, 482.2) | 639.4 (458.7, 869.8) |
|  | Sinusitis<sup>f</sup> | 7.1 (3.8, 12.0) | 31.9 (17.4, 53.5) | -- | 36.9 (20.1, 61.8) | 52.9 (28.9, 88.7) | 95.1 (51.9, 159.3) |
|  | Pneumonia (outpatient)<sup>g</sup> | 3.4 (1.9, 5.7) | 15.4 (8.8, 25.4) | -- | 17.9 (10.2, 29.4) | 25.6 (14.7, 42.2) | 46.1 (26.4, 75.7) |
|  | Pneumonia (inpatient)<sup>g</sup> | 0.3 (0.2, 0.4) | 1.2 (0.7, 1.8) | -- | 1.4 (0.8, 2.1) | 2.0 (1.2, 3.0) | 3.5 (2.1, 5.3) |
|  | IPD | 0.2 (0.2, 0.3) | 0.4 (0.4, 0.5) | -- | 0.5 (0.4, 0.6) | 0.6 (0.5, 0.7) | 0.9 (0.8, 1.0) |
| Adults ≥18 years<sup>h</sup> | Sinusitis | 162.7 (94.9, 250.2) | 731.2 (430.1, 1108.6) | 2,675.0 (1,576.4, 4,043.1) | 845.6 (497.4, 1281.4) | 1,214.0 (714.9, 1,837.6) | 2,181.8 (1,285.7, 3,298.8) |
|  | Pneumonia (outpatient)<sup>g</sup> | 46.1 (17.1, 99.3) | 109.7 (47, 205.2) | 263.9 (117.7, 451.8) | 179.9 (79.2, 318.1) | 156.9 (68.8, 279.1) | 307.9 (138, 522.4) |
|  | Pneumonia (inpatient)<sup>g</sup> | 2.0 (0.7, 4.3) | 4.9 (2.1, 9.1) | 11.8 (5.3, 20.0) | 7.9 (3.5, 13.9) | 6.9 (3.1, 12.3) | 13.8 (6.2, 23.1) |
|  | IPD | 2.9 (1.8, 3.6) | 6.1 (3.7, 7.6) | 12.3 (7.5, 15.2) | 8.2 (5, 10.2) | 9.1 (5.6, 11.3) | 13.5 (8.2, 16.7) |

Abbreviations: CI – confidence interval; PCV – pneumococcal conjugate vaccine; AOM – acute otitis media; IPD – invasive pneumococcal disease.
<sup>a</sup> PCV15 includes serotypes 1, 3, 4, 5, 6A/C, 6B, 7F, 9V, 14, 18C, 19F, 19A, 22F, 23F, 33F. PCV20 includes serotypes 1, 3, 4, 5, 6A/C, 6B, 7F, 8, 9V, 10A, 11A, 12F, 14, 15B, 18C, 19F, 19A, 22F, 23F, 33F. PCV21 includes serotypes 3, 6A/C, 7F, 8, 9N, 10A, 11A, 12F, 15A, 15C, 16F, 17F, 19A, 20A, 22F, 23A, 23B, 24F, 31, 33F, 35B. PCV24 includes serotypes 1, 2, 3, 4, 5, 6A/C, 6B, 7F, 8, 9N, 9V, 10A, 11A, 12F, 14, 15B, 17F, 18C, 19F, 19A, 20B, 22F, 23F, 33F. PCV25 includes serotypes 1, 2, 3, 4, 5, 6B, 6A/C, 7F, 8, 9N, 9V, 10A, 12F, 14, 15A, 15B, 16F, 18C, 19F, 19A, 22F, 23F, 24F, 33F, 35B. PCV31 includes serotypes 1, 2, 3, 4, 5, 6A/C, 6B, 7F, 7C, 8, 9N, 9V, 10A, 11A, 12F, 14, 15A, 15B, 16F, 17F, 18C, 19F, 19A, 20B, 22F, 23F, 23A, 23B, 31, 33F, 35B.
<sup>b</sup> Considering US population by age group<sup>33</sup> and assuming disease incidence rates as detailed in **Tables S11–S13**, pneumococcal-attributable disease proportions (**Table S4**), preventable PCV coverage proportions of pneumococci by condition and age-group (**Table S14**), PCV effectiveness (**Table S6**), and no serotype 15B and 15C cross-protection.
<sup>c</sup> Estimates for PCV21 for children 0–17 years not presented as PCV21 is not indicated for use in children.
<sup>d</sup> Also includes Pn-MAPS24v.
<sup>e</sup> Limited to children eligible for vaccination with PCV15 or PCV20: all children 0–4 years, and children 5–17 years at increased risk of pneumococcal disease (**Table S5**). PCV15 or PCV20 currently recommended for all children 2–23 months, children 24–59 months with incomplete PCV vaccination status without contraindications for vaccination, and children 5–17 years with health conditions placing them at increased risk of pneumococcal disease.<sup>9,10</sup>
<sup>f</sup> Sinusitis only considered in children 5–17 years.
<sup>g</sup> Non-bacteremic pneumonia. Bacteremic pneumonia included in IPD definition.
<sup>h</sup>Limited to adults eligible for vaccination with PCV15, PCV20, or PCV21: all adults $\geq 50$ years and adults 18–49 years at increased risk of pneumococcal disease (**Table S5**). Vaccination with PCV15 (in addition to 23-valent pneumococcal polysaccharide vaccine [PPSV23]), PCV20, or PCV21 recommended for adults 18–49 years with health conditions placing them at increased risk of pneumococcal disease.<sup>11,12</sup> Vaccination with PCV15 (plus PPSV23), PCV20, or PCV21 recommended for all adults $\geq 50$ years without contraindications for vaccination.<sup>11,12</sup>

In U.S. PCV-eligible adults, potentially preventable disease burdens included 211,888–2,496,235 outpatient-managed ARIs, 1,979–13,751 non-bacteremic pneumonia hospitalizations, and 2,913– 13,464 IPD cases annually (**Table 3**, **Table S18**). The greatest preventable burdens of adult non-bacteremic pneumonia and IPD were from PCV31. PCV21 could potentially prevent the greatest burden of sinusitis. Limited increases in adult pneumonia and IPD and large increases in sinusitis were observed with 15B/C cross-protection (**Table S19**).

## DISCUSSION

We found that serotype distribution, PCV coverage and preventable burdens varied widely by condition and age group. Overall, pneumococcal disease coverage was lowest for PCV15 and highest for PCV31. PCV20 offered nearly two-fold greater coverage of pediatric ARIs compared with PCV15; differences for IPD and adult non-bacteremic pneumonia were modest. In pediatric ARIs, PCV24 offered minimal coverage improvements over PCV20; larger increases were observed with PCV25. In adult disease, coverage from PCV24 and PCV25 were similar. Notably, PCV21 provided greater coverage than all other pipeline vaccines except PCV31 for adult pneumonia and IPD. Varying by PCV, preventable burdens encompassed 270 thousand to 3.3 million outpatient-managed ARIs, 2 to 17 thousand non-bacteremic pneumonia hospitalizations, and 3 to 14 thousand IPD cases in the United States each year.

We found that the most prevalent serotypes in pediatric pneumococcal ARIs are 15C, 23B, 11A, 15A, and 35B. No PCVs except PCV21, which is not currently indicated for pediatric use, target all these serotypes. However, PCV31 targets 23B, 11A, 15A, 35B, and 15B, with potential 15C cross-reactivity. In adult sinusitis (where serotype distribution was inferred from pediatric ARIs), PCV21 provides greater coverage than PCV31 from inclusion of 15C. The most prevalent model-estimated serotypes in adult non-bacteremic pneumonia are 3, 22F, 20B, 19A, and 35B. However, in adults ≥65 years, universally eligible for PCV immunization,^11^ 20B accounts for only 3% of pneumococcal pneumonia. All PCVs target 22F and PCV21, PCV25, PCV31 target 35B while only PCV31 targets 20B. Serotypes 22F, 23B, and 33F, common in pediatric IPD, are included in all PCV formulations. Serotypes 3 and 22F are also top contributors to adult IPD, along with 23A, 9N, and 8. PCV21 and PCV31 target 23A and PCV21, PCV24, PCV25, and PCV31 target 9N and 8.

Known as carrier suppression, increases in valency have corresponded to numerically lower serotype-specific immune responses. Thus, serotype coverage added to existing conjugate protein delivery systems may be offset by diminishing immune response. In a phase 3 trial of PCV20, immune responses one month after the third priming dose failed to meet non-inferiority criteria for five serotypes common to PCV13 and PCV20.^40^ In a phase 3 trial among adults, PCV20-elicited opsonophagocytic activity (OPA) geometric mean fold rises met non-inferiority criteria, but were numerically lower than those from PCV13 for most (11/13) shared serotypes.^41^ However, it is unknown if immune response differences translate into VE differences. Whether new protein conjugation methods (PCV24, PCV31) and MAPS technology^42^ (PN-MAPS24v) mitigate carrier suppression remains to be determined. In dose-ranging studies, immunogenicity of PCV24 delivering 2.2µg of each antigen was equal to or greater than PCV20-induced immunogenicity for shared serotypes, although carrier suppression occurred in a product delivering 4.4 µg for 7 antigens.^43,44^ Data from phase 1 and 2 trials suggest robust immune responses to Pn-MAPS24v.^45,46^ Although employing a traditional platform, PCV21 mitigates carrier suppression by dropping select serotypes targeted by existing PCVs.^47^ In estimating preventable burdens, we assume syndrome-specific VEs equivalent to PCV7/PCV13; the effects of carrier suppression on this assumption remain to be determined.

Serotype cross-protection is an important unknown. Better understanding of 15B/C cross-protection is needed to evaluate potential PCV impacts on pediatric ARIs, where 15C is prevalent. While a study of children 6–36 months found no 15B/C cross-reactivity,^48^ OPA titers against 15C were identified in a small sample (N=32) of adults 18–49 years immunized with PCV20.^26^ Additionally, a phase 3 trial demonstrated OPA responses to 15B among adults immunized with PCV21.^49^ Notably, PCV21 is indicated for prevention of serotype 15B and 15C IPD, but only for serotype 15B pneumonia,^20^ for which protection requires higher antibody levels. We assumed complete 6A/C cross-protection based on demonstrated PCV13 cross-protection in AOM and IPD.^24^

Our analysis has limitations. First, we use studies from U.S. and non-U.S. contexts to estimate pediatric ARI serotype distribution. Inclusion of non-U.S. studies from comparable contexts allowed for robust estimation. However, potential geographic variation in serotype distribution could affect our estimates.

Second, we relied on AOM studies to inform serotype distribution estimates for sinusitis and pediatric non-bacteremic pneumonia given limited available data for these conditions. Similarly, we extrapolated PCV VE against sinusitis from AOM. Third, adult non-bacteremic pneumonia serotype distribution was estimated from adults hospitalized in two major health systems in the southeastern United States and may not be nationally representative or convey to outpatient-managed pneumonia. Geographic variation is important for serotype 4, for which IPD incidence increased in 3/10 regional ABCs sites from 2010–2018^50^ and which is in all PCVs except PCV21. Fourth, serotypes causing IPD informed estimated adult non-bacteremic pneumonia serotype distribution: the PNEUMO study included bacteremic pneumococcal pneumonia cases (14.5%) and we inferred non-SSUAD serotype distribution using IPD data. Fifth, burden estimates rely on overall PCV7/PCV13 VE and do not consider serotype-specific VE, serotype replacement, or indirect effects from pediatric immunization. We assume equivalent PCV13-serotype VE for all PCVs and exclude PCV13 serotypes from preventable burden estimates, potentially underestimating preventable burdens. Underestimation may be consequential for serotype 3, which accounts for sizeable proportions of adult disease and for which PCV13 VE estimates were non-significant in pediatric IPD^22^ and adult pneumonia.^51^ Finally, post-COVID-19 pandemic healthcare utilization for pneumococcal diseases is unknown. Despite these limitations, our study provides a comprehensive analysis of best-available data to estimate the burden of pneumococcal disease in the United States preventable by PCV products across multiple conditions.

In summary, PCV21, PCV24, PCV25, and PCV31 target serotypes accounting for over 60% of IPD and adult pneumonia with lesser coverage of pediatric ARIs. Ongoing surveillance of serotype distribution in ARIs and IPD may inform future vaccine formulations and policy.

## Data Availability

Data produced in the present work are contained in the manuscript.

## ACKNOWLEDGEMENTS

The authors would like to thank Dr. Jin Han, Dr. Kelly Johnson, Dr. Thomas Weiss, Dr. Craig Roberts and all other PNEUMO Study Investigators for their contributions to the PNEUMO study, which contributed data for this analysis.

## Conflicts of Interest and Financial Disclosures

LMK reports consulting fees from Merck Sharpe & Dohme for unrelated work and Vaxcyte for related and unrelated work and reports support for attending a meeting from UC Berkeley Center for Effective Global Action. CGG reports consulting fees from Merck & Co., Inc and research funding from NIH, CDC, AHRQ, FDA, and Campbell Alliance/Syneos Health for unrelated work. NR reports research funding from Merck, Sanofi, Pfizer, Vaccine Company, Immorna, Quidel and Lilly for unrelated work, reports consulting fees from Krog, reports honoraria from Virology Education and Medscape, reports support for attending a meeting from Sanofi and Moderna, reports participation in advisory boards for Moderna, Sanofi, Seqirus, Pfizer, EMMES, ICON, and Micron, and reports leadership roles with ARLG, TMRC, CDC-Pertussis challenge, and *Clinical*

### Infectious Diseases

JAL reports research grants from Pfizer and Merck Sharpe & Dohme for unrelated work and consulting fees from Pfizer, Merck, Sharpe & Dohme, Vaxcyte, Seqirus Inc., and Valneva SE for unrelated work and Vaxcyte for related work, and reports support for attending a meeting from Vaxcyte. All other authors reported no conflicts of interest.

## Data Access, Responsibility, and Analysis

LMK had full access to all the data in the study and takes responsibility for the integrity of the data and the accuracy of the data analysis.

