## Supplemental_materials for "PNEUMOCOCCAL SEROTYPE DISTRIBUTION AND COVERAGE OF EXISTING AND PIPELINE PNEUMOCOCCAL VACCINES"

[1] School of Public Health, University of California, Berkeley, Berkeley, California, United States; [2] Division of Bacterial Diseases, Centers for Disease Control and Prevention, Atlanta, Georgia, United States; [3] Vanderbilt University Medical Center, Nashville, Tennessee, United States; [4] Rollins School of Public Health, Emory University, Atlanta, Georgia, United States; [5] Department of Medicine, Emory University, Atlanta, Georgia, United States

Corresponding author:
Laura King
University of California, Berkeley
2121 Berkeley Way
Berkeley, CA 94704


### Contents of this supplement

**Table S1.** Summary of data inputs and sources

**Table S2.** Search strings used to identify studies of serotype distribution in pediatric acute respiratory infections

**Table S3.** Studies included in meta-analysis of pneumococcal serotype distribution in nasopharyngeal samples from children with acute respiratory infection

**Table S4.** Estimates of pneumococcal-attributable (any serotype) percent of cases

**Table S5.** Percent of individuals with underlying health conditions qualifying for risk-based pneumococcal conjugate vaccines

**Table S6.** Pneumococcal conjugate vaccine effectiveness (VE) estimate inputs

**Table S7.** Estimated percent of all pneumococcal isolates belonging to each serotype

**Table S8.** Estimated percent of pneumococcal infections and number of cases potentially preventable by 21-valent pneumococcal conjugate vaccine (PCV21) in children with risk factors for pneumococcal disease

**Table S9.** Proportion of pneumococcal infections due to serotypes targeted by pneumococcal conjugate vaccine products assuming complete serotype 15B and 15C cross-protection and comparison with primary analysis estimates among children 0–17 years

**Table S10.** Proportion of pneumococcal infections due to serotypes targeted by pneumococcal conjugate vaccine products assuming complete serotype 15B and 15C cross-protection and comparison with primary analysis estimates among adults ≥18 years

**Table S11.** Estimated national burdens of invasive pneumococcal disease in 2019

**Table S12.** Estimated national burdens of all-cause AOM, pneumonia, and sinusitis visits among outpatients in 2019

**Table S13.** Estimated national burdens of all-cause pneumonia among inpatients in 2019

**Table S14.** Preventable proportion of pneumococcal infections due to serotypes targeted by pneumococcal conjugate vaccines excluding PCV13 serotypes + 6C

**Table S15.** Preventable proportion of pneumococcal infections due to serotypes targeted by pneumococcal conjugate vaccines excluding PCV13 serotypes + 6C assuming complete serotype 15B and 15C cross-protection

**Table S16.** Estimated number of cases potentially preventable by pneumococcal conjugate vaccines in children 0–17 years

**Table S17.** Estimated number of cases potentially preventable by pneumococcal conjugate vaccines assuming complete serotype 15B and 15C cross-protection in children 0–17 years

**Table S18.** Estimated number of cases potentially preventable by pneumococcal conjugate vaccines in adults ≥18 years

**Table S19.** Estimated number of cases potentially preventable by pneumococcal conjugate vaccines assuming complete serotype 15B and 15C cross-protection in adults ≥18 years

**Table S1. Summary of data inputs and sources**

| Measure | Condition | Age group | Source |
|---|---|---|---|
| Serotype distribution | AOM | Children 0–17 years | Meta-analysis of published studies (**Table S3**) |
| Serotype distribution | Non-bacteremic pneumonia | Children 0–17 years | Meta-analysis of published studies (**Table S3**)[a] |
| Serotype distribution | Sinusitis | Children and adults ≥5 years[b] | Meta-analysis of published studies (**Table S3**)[a] |
| Serotype distribution | Non-bacteremic pneumonia | Adults ≥18 years | PNEUMO study of hospitalized adults using 30-serotype SSUAD[1] + 2015–2019 ABCs[2] [c] |
| Serotype distribution | IPD | All ages | 2015–2019 ABCs[2] |
| Proportion of cases that are pneumococcal-attributable | AOM, sinusitis, non-bacteremic pneumonia | All ages | Published studies (**Table S4**) |
| Annual number and incidence of all-cause outpatient-managed visits | AOM | Children 0–17 years | 2016, 2019 NAMCS/NHAMCS + 2016–2019 MarketScan[d] |
| Annual number and incidence of all-cause outpatient-managed visits | Non-bacteremic pneumonia | Adults ≥18 years | 2016, 2019 NAMCS/NHAMCS + 2016–2019 MarketScan[d] |
| Annual number and incidence of all-cause outpatient-managed visits | Sinusitis | Children and adults ≥5 years[b] | 2016, 2019 NAMCS/NHAMCS + 2016–2019 MarketScan[d] |
| Annual number and incidence of all-cause hospitalizations | Non-bacteremic pneumonia | All ages | 2019 NIS[e] |
| Annual number and incidence of cases | IPD | All ages | 2019 ABCs |
| Proportion of cases occurring among individuals at increased risk of pneumococcal disease | Non-bacteremic pneumonia, IPD | Children and adults 5–64 years | Published studies (**Table S5**) |

| | | | |
|---|---|---|---|
| PCV effectiveness against vaccine-serotype disease | All conditions | All ages | Published studies (**Table S6**) |

Abbreviations: AOM – acute otitis media; PNEUMO -- Pneumococcal pNeumonia Epidemiology, Urine serotyping, and Mental Outcomes; SSUAD – serotype-specific urinary antigen detection; ABCs – Active Bacterial Core surveillance; IPD – invasive pneumococcal disease; NAMCS – National Ambulatory Medical Care Survey; NHAMCS – National Hospital Ambulatory Medical Care Survey; NIS – National Inpatient Sample.

[a] Based on meta-analysis of studies of serotype distribution in pediatric AOM and pneumonia.

[b] Sinusitis only considered in children ≥5 years due to incomplete sinusitis development and low counts of sinusitis diagnoses in NAMCS/NHAMCS precluding analysis in young ages.

[c] SSUAD data used for 30 SSUAD-type serotypes with additional serotype prevalence inferred from ABCs IPD data.

[d] NAMCS/NHAMCS and MarketScan methods.

[e] NIS minus bacteremic pneumonia from ABCs.

**Table S2. Search strings used to identify studies of serotype distribution in pediatric acute respiratory infections**

| String name | Search terms |
|---|---|
| **S1** | “((((*Streptococcus pneumoniae*) OR (pneumococcus)) OR (pneumococcal)) OR (*S. pneumoniae*)) AND (serotype) AND ((non-invasive) OR (outpatient))” |
| **S2** | “((((*Streptococcus pneumoniae*) OR (pneumococcus)) OR (pneumococcal)) OR (*S. pneumoniae*)) AND (serotype) AND ((otitis media) OR (otitis) OR (acute otitis media) OR (pneumonia) OR (sinusitis))” |

**Table S3. Studies included in meta-analysis of pneumococcal serotype distribution in nasopharyngeal samples from children with acute respiratory infection**

| Reference | Study years | Study country | Participant ages | Condition | Serotyping method(s) | N isolates |
|---|---|---|---|---|---|---|
| Chen 2020[3] | 2016–2018 | Taiwan | 2–60 months | AOM | Latex agglutination; Quellung reaction;[a] PCR | 42 |
| Kaur JID 2024[4] | 2011–2019 | United States | 6–36 months | AOM | Quellung reaction[a] | 400 |
| Kaur PIDJ 2024[5] | 2021–2023 | United States | 6–36 months | AOM | Quellung reaction[a] | 79 |
| Lewnard 2021[6,b] | 2009–2018 | Israel | 0–59 months | Pneumonia | Quellung reaction[a] | 117 |
| Martin 2018[7] | 2012–2014 | United States | 6–23 months | AOM | Quellung reaction[a] | 113 |
| Rybak 2023[8] | 2010–2022 | France | 6–24 months | AOM | Latex agglutination | 5759 |

Abbreviations: N – number; AOM – acute otitis media; PCR – polymerase chain reaction; JID – Journal of Infectious Diseases; PIDJ – Pediatric Infectious Diseases Journal.

[a] Serum Staten Institute, Copenhagen, Denmark.

[b] Only samples from Jewish children included (Bedouin children excluded). Although a small proportion of samples are from the pre-PCV13 period, we included this study as few others with serotype-specific estimates were available for pneumonia or other lower respiratory tract infections.

**Table S4. Estimates of pneumococcal-attributable (any serotype) percent of cases**

| Condition, age group | Reported percent of cases attributable to *S. pneumoniae* (95% CI) | Source |
|---|---|---|
| AOM, children 0–17 years | 19.4 (16.8, 22.2) | King 2024[9] |
| Sinusitis, children 0–17 years | 45.7 (28.4, 63.7) | King 2024[9] |
| Non-bacteremic pneumonia, children 0–17 years | 27.1 (17.4, 38.5) | King 2024[9] |
| Sinusitis, adults ≥18 years[b] | 45.7 (28.4, 63.7) | King 2024[9] |
| Non-bacteremic pneumonia,[c] adults 18–49 years | 9.9 (8.1, 12.1) | Self 2024[1] |
| Non-bacteremic pneumonia,[c] adults 50–64 years | 14.7 (12.5, 17.0) | Self 2024[1] |
| Non-bacteremic pneumonia,[c] adults ≥65 years | 11.4 (9.6, 13.3) | Self 2024[1] |

Abbreviations: CI – confidence interval; AOM – acute otitis media.

[a] Estimates presented are those reported. To propagate uncertainty when including multiple inputs in estimates of preventable burdens, we fit the reported pneumococcal-attributable percent values to gamma distributions and sampled 4,900,000 draws from these distributions.

[b] The percent of adult sinusitis due to pneumococcus assumed to be consistent with the pneumococcal-attributable percent in pediatric sinusitis.

[c] Original study included both bacteremic and non-bacteremic pneumonia cases in their case definition.

**Table S5. Percent of individuals with underlying health conditions qualifying for risk-based pneumococcal conjugate vaccines[a]**

| Condition | Setting | Age group | Estimated percent (95% CI)[b] | Source |
|---|---|---|---|---|
| Pneumococcal pneumonia[c] | | | | |
| | All | 5–17 years | 21.6 (20.3, 23.0) | Pelton 2014[10] |
| | Outpatient[d] | 18–49 years | 43.8 (43.0, 44.5) | Grant 2023[11] |
| | Inpatient[d] | 18–49 years | 58.5 (57.4, 59.5) | Grant 2023[11] |
| IPD | | | | |
| | All | 5–17 years | 31.0 (25.9, 36.3) | Pelton 2014[10] |
| | All | 18–49 years | 68.3 (63.8, 72.5) | Grant 2023[11] |

Abbreviations: PCV – pneumococcal conjugate vaccine; CI – confidence interval; IPD – invasive pneumococcal disease.
[a] PCV15, PCV20, or PCV21 in adults and PCV15 or PCV20 in children.[12–15]
[b] Obtained by fitting the no. cases among individuals with underlying health conditions / total no. cases and corresponding standard error to beta distributions and sampling 4,900,000 draws.
[c] Pneumococcal pneumonia estimates applied to all respiratory infections (acute otitis media, sinusitis) in corresponding age groups.
[d] Estimated using the proportion of cases occurring in each setting for all adults 18–64 years (Grant 2023[11] Supplemental Table 6) and number of cases in each age group (Grant 2023[11] Supplemental Table 5).

**Table S6. Pneumococcal conjugate vaccine effectiveness (VE) estimate inputs[a]**

| Condition | PCV | Reported PCV effectiveness, % (95% CI)[a,b] | Source |
|---|---|---|---|
| Pediatric AOM | PCV7 | 57.0 (44.0, 67.0) | Eskola 2001[16] |
| Pediatric sinusitis[c] | PCV7 | 57.0 (44.0, 67.0) | Eskola 2001[16] |
| Pediatric non-bacteremic pneumonia | PCV13 | 53.2 (45.4, 57.2) | Estimated[c] |
| Pediatric IPD | PCV13 | 90.2 (75.4, 96.1) | Andrejko 2024[17] |
| Adult sinusitis[c] | PCV7 | 57.0 (44.0, 67.0) | Eskola 2001[16] |
| Adult non-bacteremic pneumonia | PCV13 | 45.0 (14.2, 65.3) | Bonten 2015[18] |
| Adult IPD | PCV13 | 75.0 (41.4, 90.8) | Bonten 2015[18] |

Abbreviations: CI – confidence interval; PCV – pneumococcal conjugate vaccine; AOM – acute otitis media; IPD – invasive pneumococcal disease.

[a] Estimates presented are those reported in previous studies. To propagate uncertainty when including multiple inputs in estimates of preventable burdens, we fit the reported VE values to beta distributions and sampled 4,900,000 draws from these distributions.

[b] VE against vaccine-type disease.

[c] Sinusitis VE considered to be consistent with that for AOM; no studies of PCV VE against sinusitis are available.

[d] Obtained by applying the ratio of PCV13 VE in adult CAP to adult IPD (45:75) to PCV13 VE against pediatric IPD estimates consistent with prior studies.[19]

**Table S7. Estimated percent of all pneumococcal isolates belonging to each serotype**

| | Estimated percent of all pneumococcal isolates (95% CI)[a] | | | | | | | | |
|---|---|---|---|---|---|---|---|---|---|
| **Sero-type** | *AOM 0–17 years* | *Pneumonia,[b] sinusitis 0–17 years* | *Pneumonia[b] 18–49 years* | *Pneumonia[b] 50–64 years* | *Pneumonia[b] ≥65 years* | *IPD 0–17 years* | *IPD 18–49 years* | *IPD 50–64 years* | *IPD ≥65 years* |
| **23B** | 9.4 (8.7, 10.2) | 9.3 (8.6, 10.0) | 6.0 (2.4, 12.4) | 2.2 (0.6, 5.3) | 1.7 (0.3, 4.8) | 7.7 (6.2, 9.4) | 3.4 (2.7, 4.1) | 3.3 (2.8, 3.9) | 2.9 (2.5, 3.4) |
| **11A** | 9.0 (8.3, 9.7) | 8.8 (8.2, 9.6) | 2.0 (0.4, 6.6) | 4.3 (2.0, 8.4) | 4.0 (1.8, 8.3) | 2.1 (1.4, 3.2) | 3.1 (2.5, 3.8) | 3.7 (3.2, 4.3) | 4.2 (3.7, 4.7) |
| **15A** | 7.8 (7.2, 8.5) | 7.8 (7.1, 8.4) | 0 (0, 0) | 2.7 (0.8, 6.1) | 0 (0, 0) | 4.2 (3.1, 5.5) | 2.8 (2.2, 3.5) | 3.9 (3.4, 4.5) | 5.7 (5.1, 6.3) |
| **35B** | 7.7 (7.0, 8.3) | 7.6 (6.9, 8.2) | 7.4 (3.0, 14.3) | 3.6 (1.3, 7.4) | 3.9 (1.4, 8.1) | 5.3 (4.0, 6.7) | 3.0 (2.4, 3.7) | 3.5 (3, 4) | 6.1 (5.5, 6.8) |
| **23A** | 5.9 (5.4, 6.5) | 5.9 (5.3, 6.5) | 4.2 (1.3, 9.4) | 2.4 (0.7, 5.5) | 5.5 (2.6, 9.9) | 2.7 (1.9, 3.8) | 3.9 (3.2, 4.7) | 4.5 (3.9, 5.1) | 6.0 (5.4, 6.6) |
| **21** | 5 (4.5, 5.5) | 4.9 (4.4, 5.5) | 0 (0, 2.0) | 0.4 (0.1, 2.6) | 0 (0, 1.3) | 2.6 (1.8, 3.7) | 0.2 (0.1, 0.5) | 0.2 (0.1, 0.4) | 0.2 (0.1, 0.3) |
| **19A** | 4.3 (3.8, 4.8) | 4.3 (3.8, 4.8) | 8.7 (3.8, 15.7) | 5 (2.2, 9.2) | 4.1 (1.5, 8.4) | 6.4 (5, 8) | 3.7 (3, 4.4) | 3.7 (3.2, 4.2) | 2.9 (2.5, 3.3) |
| **35F** | 4.3 (3.8, 4.8) | 4.2 (3.8, 4.8) | 0.9 (0, 4.2) | 2.4 (0.6, 6.0) | 2 (0.3, 5.4) | 1.7 (1, 2.6) | 0.9 (0.6, 1.3) | 1.5 (1.1, 1.8) | 1.5 (1.2, 1.9) |
| **10A** | 4.0 (3.5, 4.5) | 4.0 (3.5, 4.5) | 0 (0, 0.1) | 0 (0, 0.1) | 0.5 (0, 2.2) | 3.4 (2.4, 4.5) | 1.8 (1.4, 2.3) | 2.0 (1.6, 2.4) | 1.4 (1.1, 1.7) |
| **24F** | 3.0 (2.6, 3.4) | 2.9 (2.5, 3.4) | 0.4 (0, 2.7) | 0.9 (0.1, 3.0) | 0 (0, 0) | 0 (0, 0) | 0 (0, 0) | 0 (0, 0) | 0 (0, 0) |
| **19F** | 2.9 (2.5, 3.3) | 2.9 (2.5, 3.4) | 0.4 (0, 3.5) | 5.4 (2.5, 9.5) | 4.6 (1.8, 8.7) | 7.7 (6.2, 9.4) | 3.4 (2.7, 4.1) | 3.0 (2.6, 3.5) | 2.4 (2.0, 2.8) |
| **NT** | 2.3 (2.0, 2.7) | 2.4 (2.1, 2.8) | 0.1 (0, 1.6) | 0 (0, 1.0) | 0 (0, 1.2) | 0.3 (0.1, 0.7) | 0.1 (0, 0.2) | 0.1 (0.1, 0.2) | 0.1 (0.0, 0.2) |
| **22F** | 2.3 (2.0, 2.7) | 2.3 (1.9, 2.7) | 7.2 (2.7, 13.7) | 9.9 (5.8, 15.3) | 5.3 (2.1, 9.9) | 8.9 (7.3, 10.7) | 9.4 (8.3, 10.5) | 10.3 (9.4, 11.2) | 10.9 (10.1, 11.7) |
| **6C[c]** | 2.3 (1.9, 2.7) | 2.3 (1.9, 2.6) | - | - | - | 1.0 (0.5, 1.7) | 2.6 (2.1, 3.3) | 2.4 (2.0, 2.9) | 3.7 (3.3, 4.3) |
| **15C** | 11.4 (10.6, 12.3) | 11.4 (10.5, 12.2) | 1.6 (0.3, 5.5) | 0.3 (0, 2.1) | 1.7 (0.5, 4.6) | 5.9 (4.6, 7.4) | 0.9 (0.6, 1.3) | 0.8 (0.6, 1.1) | 1.2 (1.0, 1.5) |
| **33F** | 1.9 (1.6, 2.2) | 1.9 (1.5, 2.2) | 0.6 (0, 4.1) | 1 (0.1, 3.5) | 0.4 (0, 2.7) | 6.7 (5.3, 8.3) | 3.8 (3.1, 4.5) | 3.3 (2.8, 3.8) | 4.4 (3.9, 5.0) |
| **31** | 1.7 (1.4, 2.0) | 1.7 (1.4, 2.0) | 0 (0, 0) | 4.0 (1.6, 8.2) | 2.3 (0.6, 6.0) | 0.6 (0.2, 1.2) | 1.6 (1.2, 2.1) | 1.9 (1.6, 2.4) | 2.8 (2.4, 3.3) |
| **17F** | 1.7 (1.4, 2.0) | 1.6 (1.3, 2.0) | 3.0 (0.5, 7.9) | 1.7 (0.3, 4.6) | 3.3 (1.0, 7.2) | 0.6 (0.3, 1.3) | 1.7 (1.2, 2.2) | 1.3 (1.0, 1.6) | 1.3 (1.0, 1.6) |
| **16F** | 1.6 (1.3, 1.9) | 1.6 (1.3, 2.0) | 1.3 (0.2, 5.8) | 0.2 (0, 1.9) | 3.7 (1.4, 7.9) | 3.1 (2.1, 4.2) | 3.0 (2.4, 3.7) | 3.8 (3.2, 4.3) | 4.4 (3.9, 5.0) |

| | Estimated percent of all pneumococcal isolates (95% CI)[a] | | | | | | | | |
|---|---|---|---|---|---|---|---|---|---|
| **Sero-type** | *AOM 0–17 years* | *Pneumonia,[b] sinusitis 0–17 years* | *Pneumonia[b] 18–49 years* | *Pneumonia[b] 50–64 years* | *Pneumonia[b] ≥65 years* | *IPD 0–17 years* | *IPD 18–49 years* | *IPD 50–64 years* | *IPD ≥65 years* |
| **15B** | 1.6 (1.3, 1.9) | 1.5 (1.2, 1.9) | 1.7 (0.3, 5.8) | 1.7 (0.4, 4.4) | 3.2 (1.1, 6.9) | 3.9 (2.9, 5.2) | 1.3 (1.0, 1.8) | 1.1 (0.8, 1.4) | 2.0 (1.7, 2.4) |
| **3** | 1.4 (1.1, 1.7) | 1.4 (1.1, 1.7) | 9.3 (4.7, 16.5) | 13.0 (8.6, 18.8) | 10.8 (6.6, 16.6) | 9.0 (7.4, 10.9) | 11.2 (10.1, 12.4) | 14.3 (13.3, 15.3) | 15.8 (14.9, 16.8) |
| **9N** | 1.2 (0.9, 1.5) | 1.2 (1.0, 1.5) | 6.5 (2.5, 12.9) | 5.6 (2.7, 10.0) | 1.9 (0.4, 5.4) | 1.9 (1.2, 2.8) | 7.4 (6.5, 8.4) | 6.9 (6.2, 7.6) | 4.0 (3.5, 4.6) |
| **29** | 0.9 (0.7, 1.1) | 0.9 (0.7, 1.1) | 0 (0, 0.9) | 0 (0, 0.7) | 0 (0, 0.7) | 0 (0, 0.1) | 0 (0, 0) | 0 (0, 0) | 0 (0, 0) |
| **11C** | 0.9 (0.5, 1.5) | 0.9 (0.5, 1.4) | 0 (0, 1.0) | 0 (0, 0.8) | 0 (0, 0.7) | 0 (0, 0.1) | 0 (0, 0) | 0 (0, 0) | 0 (0, 0) |
| **35D** | 0.7 (0.4, 1.3) | 0.7 (0.4, 1.3) | 0 (0, 0.1) | 0 (0, 0.1) | 0 (0, 0.1) | 0.1 (0, 0.4) | 0 (0, 0.1) | 0.1 (0, 0.2) | 0.1 (0, 0.2) |
| **6A[c]** | 0.6 (0.4, 0.8) | 0.4 (0.2, 0.6) | 2.0 (0.3, 5.9) | 0.6 (0.1, 2.5) | 4.8 (1.9, 8.9) | 0 (0, 0.1) | 0.1 (0, 0.3) | 0.1 (0.1, 0.3) | 0.1 (0.1, 0.2) |
| **40** | 0.4 (0.2, 1) | 0.4 (0.2, 0.9) | 0 (0, 0.6) | 0 (0, 0.5) | 0 (0, 0.5) | 0 (0, 0.1) | 0 (0, 0) | 0 (0, 0) | 0 (0, 0) |
| **11D** | 0.3 (0.1, 0.7) | 0.3 (0.1, 0.7) | 0 (0, 0.8) | 0 (0, 0.5) | 0 (0, 0.6) | 0 (0, 0.1) | 0 (0, 0) | 0 (0, 0) | 0 (0, 0) |
| **34** | 0.3 (0.1, 0.7) | 0.3 (0.1, 0.6) | 1.5 (0.3, 6.1) | 1.5 (0.4, 4.6) | 2.4 (0.7, 6.2) | 0.9 (0.4, 1.6) | 1.1 (0.8, 1.6) | 1.3 (1.0, 1.6) | 1.6 (1.3, 2.0) |
| **38** | 0.3 (0, 0.7) | 0.3 (0, 0.7) | 0.3 (0, 4.1) | 1.4 (0.2, 4.5) | 1.5 (0.3, 4.9) | 2.4 (1.6, 3.4) | 0.7 (0.4, 1.0) | 1.0 (0.8, 1.4) | 1.4 (1.1, 1.8) |
| **23F** | 0.2 (0.1, 0.4) | 0.1 (0, 0.2) | 0 (0, 0.1) | 0 (0, 0.1) | 0 (0, 0.1) | 0 (0, 0.3) | 0.1 (0, 0.2) | 0.1 (0, 0.1) | 0 (0, 0.1) |
| **7C** | 0.2 (0, 0.6) | 0.2 (0, 0.6) | 1.8 (0.5, 5.3) | 2.3 (0.9, 5.3) | 3.2 (1.4, 6.7) | 2.0 (1.2, 2.9) | 1.6 (1.2, 2.1) | 1.5 (1.2, 1.9) | 2.4 (2.0, 2.8) |
| **13** | 0.2 (0, 0.6) | 0.2 (0, 0.5) | 0.8 (0.1, 3.9) | 0.5 (0, 2.2) | 0 (0, 0.8) | 0.1 (0, 0.3) | 0.4 (0.2, 0.7) | 0.2 (0.1, 0.3) | 0.1 (0.1, 0.2) |
| **25A** | 0.1 (0, 0.5) | 0.1 (0, 0.5) | 0 (0, 0) | 0 (0, 0) | 0 (0, 0) | 0 (0, 0) | 0 (0, 0) | 0 (0, 0) | 0 (0, 0) |
| **28F** | 0.1 (0, 0.5) | 0.1 (0, 0.5) | 0 (0, 0) | 0 (0, 0) | 0 (0, 0) | 0 (0, 0) | 0 (0, 0) | 0 (0, 0) | 0 (0, 0) |
| **37** | 0.1 (0, 0.5) | 0.1 (0, 0.4) | 0 (0, 0.1) | 0 (0, 0.1) | 0 (0, 0.1) | 0 (0, 0) | 0.1 (0, 0.3) | 0.1 (0, 0.2) | 0.1 (0, 0.2) |
| **7A** | 0.1 (0, 0.5) | 0.1 (0, 0.4) | 0 (0, 1.0) | 0 (0, 0.5) | 0 (0, 0.7) | 0 (0, 0.1) | 0 (0, 0) | 0 (0, 0) | 0 (0, 0) |
| **15F** | 0.1 (0, 0.4) | 0.1 (0, 0.4) | 0 (0, 0.9) | 0 (0, 0.5) | 0 (0, 0.5) | 0 (0, 0.4) | 0 (0, 0.1) | 0 (0, 0) | 0 (0, 0) |
| **19B** | 0.1 (0, 0.4) | 0.1 (0, 0.4) | 0 (0, 0.3) | 0 (0, 0.2) | 0 (0, 0.2) | 0 (0, 0.1) | 0 (0, 0) | 0 (0, 0) | 0 (0, 0) |
| **33D** | 0.1 (0, 0.4) | 0.1 (0, 0.3) | 0 (0, 0.9) | 0 (0, 0.7) | 0 (0, 0.8) | 0 (0, 0.1) | 0 (0, 0.1) | 0 (0, 0) | 0 (0, 0) |
| **39** | 0.1 (0, 0.3) | 0.1 (0, 0.3) | 0 (0, 0.1) | 0 (0, 0.4) | 0 (0, 0.1) | 0 (0, 0) | 0 (0, 0) | 0 (0, 0) | 0 (0, 0) |

| Sero-type | Estimated percent of all pneumococcal isolates (95% CI)[a] | | | | | | | | |
|---|---|---|---|---|---|---|---|---|---|
| | *AOM 0–17 years* | *Pneumonia,[b] sinusitis 0–17 years* | *Pneumonia[b] 18–49 years* | *Pneumonia [b] 50–64 years* | *Pneumonia [b] ≥65 years* | *IPD 0–17 years* | *IPD 18–49 years* | *IPD 50–64 years* | *IPD ≥65 years* |
| **4** | 0.1 (0, 0.3) | 0 (0, 0.1) | 0 (0, 0.4) | 1.1 (0.1, 3.1) | 0.4 (0, 2.1) | 0.3 (0.1, 0.8) | 5.6 (4.7, 6.4) | 2.7 (2.3, 3.2) | 0.6 (0.4, 0.9) |
| **14** | 0 (0, 0) | 0.4 (0.3, 0.6) | 0 (0, 1.8) | 0.3 (0, 2.7) | 0 (0, 1.2) | 0 (0, 0.2) | 0.1 (0, 0.2) | 0.1 (0, 0.2) | 0.2 (0.1, 0.3) |
| **12F** | 0 (0, 0) | 0.1 (0, 0.3) | 0.6 (0.1, 3.4) | 0 (0, 0.7) | 0 (0, 0.8) | 2.6 (1.8, 3.7) | 6.0 (5.2, 6.9) | 5.0 (4.4, 5.7) | 2.3 (1.9, 2.7) |
| **7F** | 0 (0, 0) | 0.1 (0, 0.3) | 0.2 (0, 4.3) | 2.0 (0.6, 5.4) | 3.7 (1.4, 7.9) | 0.7 (0.3, 1.3) | 2.5 (1.9, 3.1) | 1.6 (1.2, 2.0) | 0.6 (0.5, 0.9) |
| **10C** | 0 (0, 0) | 0 (0, 0) | 0 (0, 0.4) | 0 (0, 0.2) | 0 (0, 0.3) | 0 (0, 0) | 0 (0, 0) | 0 (0, 0) | 0 (0, 0) |
| **10F** | 0 (0, 0) | 0 (0, 0) | 0 (0, 1.0) | 0 (0, 0.6) | 0 (0, 0.6) | 0.2 (0, 0.5) | 0 (0, 0.1) | 0.1 (0, 0.2) | 0.1 (0, 0.2) |
| **11B** | 0 (0, 0) | 0 (0, 0) | 0 (0, 1.2) | 0 (0, 0.9) | 0 (0, 1.0) | 0 (0, 0.2) | 0 (0, 0.1) | 0 (0, 0) | 0 (0, 0) |
| **11E** | 0 (0, 0) | 0 (0, 0) | 0 (0, 0.5) | 0 (0, 0.4) | 0 (0, 0.3) | 0 (0, 0.1) | 0 (0, 0) | 0 (0, 0) | 0 (0, 0) |
| **12A** | 0 (0, 0) | 0 (0, 0) | 0 (0, 0) | 0 (0, 0) | 0 (0, 0) | 0 (0, 0) | 0 (0, 0) | 0 (0, 0) | 0 (0, 0) |
| **12B** | 0 (0, 0) | 0 (0, 0) | 0 (0, 0.3) | 0 (0, 0.3) | 0 (0, 0.2) | 0 (0, 0) | 0 (0, 0) | 0 (0, 0) | 0 (0, 0) |
| **16A** | 0 (0, 0) | 0 (0, 0) | 0 (0, 0) | 0 (0, 0) | 0 (0, 0) | 0 (0, 0) | 0 (0, 0) | 0 (0, 0) | 0 (0, 0) |
| **17A** | 0 (0, 0) | 0 (0, 0) | 0 (0, 1.8) | 0 (0, 1.2) | 0 (0, 1.1) | 0 (0, 0.2) | 0 (0, 0.1) | 0 (0, 0) | 0 (0, 0) |
| **18A** | 0 (0, 0) | 0 (0, 0) | 0 (0, 0) | 0 (0, 0) | 0 (0, 0) | 0 (0, 0) | 0 (0, 0.2) | 0 (0, 0.1) | 0 (0, 0) |
| **18F** | 0 (0, 0) | 0 (0, 0) | 0 (0, 1.5) | 0 (0, 1.0) | 0 (0, 1.1) | 0 (0, 0.1) | 0 (0, 0.1) | 0 (0, 0) | 0 (0, 0) |
| **19C** | 0 (0, 0) | 0 (0, 0) | 0 (0, 0) | 0 (0, 0) | 0 (0, 0) | 0 (0, 0) | 0 (0, 0) | 0 (0, 0) | 0 (0, 0) |
| **27** | 0 (0, 0) | 0 (0, 0) | 0 (0, 0) | 0 (0, 0) | 0 (0, 0) | 0 (0, 0) | 0 (0, 0) | 0 (0, 0) | 0 (0, 0) |
| **28A** | 0 (0, 0) | 0 (0, 0) | 0 (0, 0.1) | 0.2 (0, 1.8) | 0.2 (0, 1.9) | 0 (0, 0.3) | 0.2 (0.1, 0.3) | 0.2 (0.1, 0.4) | 0.3 (0.2, 0.4) |
| **32A** | 0 (0, 0) | 0 (0, 0) | 0 (0, 0.2) | 0 (0, 0.3) | 0 (0, 0.4) | 0 (0, 0) | 0 (0, 0) | 0 (0, 0) | 0 (0, 0) |
| **33A** | 0 (0, 0) | 0 (0, 0) | 0 (0, 0.1) | 0 (0, 0.1) | 0 (0, 0.1) | 0.1 (0, 0.3) | 0 (0, 0.1) | 0 (0, 0) | 0 (0, 0) |
| **35A** | 0 (0, 0) | 0 (0, 0) | 0 (0, 0.2) | 0 (0, 0.2) | 0 (0, 0.1) | 0 (0, 0) | 0 (0, 0) | 0 (0, 0) | 0 (0, 0) |
| **36** | 0 (0, 0) | 0 (0, 0) | 0 (0, 0) | 0 (0, 0) | 0 (0, 0) | 0 (0, 0) | 0 (0, 0) | 0 (0, 0) | 0 (0, 0) |
| **47F** | 0 (0, 0) | 0 (0, 0) | 0 (0, 0.7) | 0 (0, 0.3) | 0 (0, 0.4) | 0 (0, 0) | 0 (0, 0) | 0 (0, 0) | 0 (0, 0) |
| **6D** | 0 (0, 0) | 0 (0, 0) | 0 (0, 0.6) | 0 (0, 0.4) | 0 (0, 0.3) | 0 (0, 0.1) | 0 (0, 0) | 0 (0, 0) | 0 (0, 0.1) |
| **6H** | 0 (0, 0) | 0 (0, 0) | 0.1 (0, 2.1) | 0.1 (0, 1.1) | 0.1 (0, 1.2) | 0 (0, 0.2) | 0 (0, 0.1) | 0 (0, 0) | 0 (0, 0) |
| **8** | 0 (0, 0) | 0 (0, 0) | 2.7 (0.6, 7.4) | 1.6 (0.4, 4.3) | 3.1 (1.1, 6.9) | 1.9 (1.2, 2.9) | 7 (6.1, 8.0) | 5.2 (4.6, 5.9) | 2.9 (2.5, 3.4) |

| | Estimated percent of all pneumococcal isolates (95% CI)[a] | | | | | | | | |
|---|---|---|---|---|---|---|---|---|---|
| **Sero-type** | *AOM 0–17 years* | *Pneumonia,[b] sinusitis 0–17 years* | *Pneumonia[b] 18–49 years* | *Pneumonia[b] 50–64 years* | *Pneumonia[b] ≥65 years* | *IPD 0–17 years* | *IPD 18–49 years* | *IPD 50–64 years* | *IPD ≥65 years* |
| **24A** | 0 (0, 0) | 0 (0, 0.1) | 0 (0, 0.1) | 0 (0, 0.1) | 0 (0, 0.1) | 0 (0, 0) | 0 (0, 0) | 0 (0, 0) | 0 (0, 0) |
| **20A** | 0 (0, 0.3) | 0 (0, 0.3) | 2.5 (0.6, 7.2) | 2.1 (0.7, 5.2) | 0.9 (0.1, 3.4) | 0 (0, 0) | 0 (0, 0) | 0 (0, 0) | 0 (0, 0) |
| **44** | 0 (0, 0.3) | 0 (0, 0.3) | 0 (0, 0) | 0 (0, 0) | 0 (0, 0) | 0 (0, 0) | 0 (0, 0) | 0 (0, 0) | 0 (0, 0) |
| **48** | 0 (0, 0.3) | 0 (0, 0.3) | 0 (0, 1.1) | 0 (0, 0.7) | 0 (0, 0.6) | 0 (0, 0.1) | 0 (0, 0) | 0 (0, 0) | 0 (0, 0) |
| **9L** | 0 (0, 0.3) | 0 (0, 0.3) | 0 (0, 0.9) | 0 (0, 0.7) | 0 (0, 0.6) | 0 (0, 0.1) | 0 (0, 0) | 0 (0, 0.1) | 0 (0, 0.1) |
| **33C** | 0 (0, 0.3) | 0 (0, 0.2) | 0 (0, 0.3) | 0 (0, 0.2) | 0 (0, 0.2) | 0 (0, 0.1) | 0 (0, 0) | 0 (0, 0) | 0 (0, 0) |
| **6B** | 0 (0, 0.2) | 0.1 (0, 0.2) | 0 (0, 0.8) | 1.1 (0.1, 3.5) | 0.5 (0, 2.6) | 0.2 (0, 0.5) | 0.1 (0, 0.2) | 0.2 (0.1, 0.3) | 0.2 (0.1, 0.4) |
| **9V** | 0 (0, 0.2) | 0.1 (0, 0.2) | 2.9 (0.7, 7.1) | 0.4 (0, 2) | 1.1 (0.2, 3.4) | 0.1 (0, 0.3) | 0.1 (0, 0.3) | 0.3 (0.1, 0.4) | 0.1 (0, 0.2) |
| **10D** | 0 (0, 0.2) | 0 (0, 0.2) | 0 (0, 2.5) | 0 (0, 1.5) | 0 (0, 1.5) | 0 (0, 0.2) | 0 (0, 0.1) | 0 (0, 0.1) | 0 (0, 0) |
| **20B** | 0 (0, 0.2) | 0 (0, 0.2) | 7.2 (3, 13.9) | 6.7 (3.4, 11.3) | 3.1 (1, 7) | 0.8 (0.4, 1.5) | 4.3 (3.6, 5.1) | 4.4 (3.8, 5) | 2.4 (2, 2.8) |
| **25F** | 0 (0, 0.2) | 0 (0, 0.2) | 0 (0, 0.4) | 0 (0, 0.2) | 0 (0, 0.3) | 0 (0, 0) | 0 (0, 0) | 0 (0, 0) | 0 (0, 0) |
| **32F** | 0 (0, 0.2) | 0 (0, 0.2) | 0 (0, 1.2) | 0 (0, 0.9) | 0 (0, 0.9) | 0 (0, 0.1) | 0 (0, 0) | 0 (0, 0) | 0 (0, 0) |
| **33B** | 0 (0, 0.2) | 0 (0, 0.2) | 0 (0, 1) | 0 (0, 0.5) | 0 (0, 0.5) | 0 (0, 0.1) | 0 (0, 0) | 0 (0, 0) | 0 (0, 0) |
| **35C** | 0 (0, 0.2) | 0 (0, 0.2) | 0 (0, 0.6) | 0 (0, 0.5) | 0 (0, 0.4) | 0 (0, 0.1) | 0 (0, 0) | 0 (0, 0) | 0 (0, 0) |
| **42** | 0 (0, 0.2) | 0 (0, 0.2) | 0 (0, 0.2) | 0 (0, 0.1) | 0 (0, 0.1) | 0 (0, 0) | 0 (0, 0) | 0 (0, 0) | 0 (0, 0) |
| **7D** | 0 (0, 0.2) | 0 (0, 0.2) | 0 (0, 0) | 0 (0, 0) | 0 (0, 0) | 0 (0, 0) | 0 (0, 0) | 0 (0, 0) | 0 (0, 0) |
| **9A** | 0 (0, 0.2) | 0 (0, 0.2) | 0 (0, 0.4) | 0 (0, 0.2) | 0 (0, 0.2) | 0 (0, 0) | 0 (0, 0) | 0 (0, 0) | 0 (0, 0) |
| **24C** | 0 (0, 0.2) | 0 (0, 0.1) | 0 (0, 0.3) | 0 (0, 0.2) | 0 (0, 0.2) | 0 (0, 0) | 0 (0, 0) | 0 (0, 0) | 0 (0, 0) |
| **43** | 0 (0, 0.2) | 0 (0, 0.1) | 0 (0, 1.2) | 0 (0, 0.8) | 0 (0, 1) | 0 (0, 0.1) | 0 (0, 0.1) | 0 (0, 0) | 0 (0, 0) |
| **1** | 0 (0, 0.1) | 0.3 (0.1, 0.5) | 0 (0, 0.7) | 0.5 (0, 2.3) | 4 (1.4, 8.3) | 0 (0, 0.1) | 0.1 (0, 0.3) | 0 (0, 0) | 0 (0, 0) |
| **5** | 0 (0, 0.1) | 0.1 (0, 0.2) | 1.7 (0.5, 5.2) | 1 (0.3, 3) | 1.8 (0.7, 4.4) | 0 (0, 0) | 0 (0, 0) | 0 (0, 0.1) | 0 (0, 0) |
| **18C** | 0 (0, 0.1) | 0 (0, 0) | 0 (0, 1.3) | 0 (0, 0.9) | 0 (0, 0.9) | 0.3 (0.1, 0.9) | 0.1 (0, 0.2) | 0.1 (0, 0.2) | 0 (0, 0) |
| **10B** | 0 (0, 0.1) | 0 (0, 0.4) | 0 (0, 0) | 0 (0, 0) | 0 (0, 0) | 0 (0, 0) | 0 (0, 0) | 0 (0, 0) | 0 (0, 0) |
| **11F** | 0 (0, 0.1) | 0 (0, 0.1) | 0 (0, 0.9) | 0 (0, 0.7) | 0 (0, 0.6) | 0 (0, 0.1) | 0 (0, 0) | 0 (0, 0) | 0 (0, 0) |
| **18B** | 0 (0, 0.1) | 0 (0, 0.1) | 0 (0, 0.1) | 0 (0, 0.1) | 0 (0, 0.1) | 0 (0, 0) | 0 (0, 0) | 0 (0, 0) | 0 (0, 0.1) |

| | Estimated percent of all pneumococcal isolates (95% CI)[a] | | | | | | | | |
|---|---|---|---|---|---|---|---|---|---|
| **Sero-type** | *AOM 0–17 years* | *Pneumonia,[b] sinusitis 0–17 years* | *Pneumonia[b] 18–49 years* | *Pneumonia[b] 50–64 years* | *Pneumonia[b] ≥65 years* | *IPD 0–17 years* | *IPD 18–49 years* | *IPD 50–64 years* | *IPD ≥65 years* |
| **2** | 0.0 (0, 0.1) | 0 (0, 0.1) | 0 (0, 0.2) | 0 (0, 0.2) | 0 (0, 0.3) | 0 (0, 0) | 0 (0, 0) | 0 (0, 0) | 0 (0, 0) |
| **22A** | 0 (0, 0.1) | 0 (0, 0.1) | 0 (0, 0.1) | 0 (0, 0.1) | 0 (0, 0.1) | 0 (0, 0) | 0 (0, 0) | 0 (0, 0) | 0 (0, 0) |
| **24B** | 0 (0, 0.1) | 0 (0, 0.1) | 0.1 (0, 1.2) | 0.1 (0, 0.8) | 0.1 (0, 0.8) | 0 (0, 0.1) | 0 (0, 0) | 0 (0, 0) | 0 (0, 0) |
| **41A** | 0 (0, 0.1) | 0 (0, 0.1) | 0 (0, 1.8) | 0 (0, 1.2) | 0 (0, 1.3) | 0 (0, 0.2) | 0 (0, 0.1) | 0 (0, 0) | 0 (0, 0) |
| **41F** | 0 (0, 0.1) | 0 (0, 0.1) | 0 (0, 0.6) | 0 (0, 0.3) | 0 (0, 0.4) | 0 (0, 0.1) | 0 (0, 0) | 0 (0, 0) | 0 (0, 0) |
| **45** | 0 (0, 0.1) | 0 (0, 0.1) | 0 (0, 0.2) | 0 (0, 0.2) | 0 (0, 0.2) | 0 (0, 0) | 0 (0, 0) | 0 (0, 0) | 0 (0, 0.1) |
| **46** | 0 (0, 0.1) | 0 (0, 0.1) | 0 (0, 0) | 0 (0, 0) | 0 (0, 0) | 0 (0, 0) | 0 (0, 0) | 0 (0, 0) | 0 (0, 0) |
| **47A** | 0 (0, 0.1) | 0 (0, 0.1) | 0 (0, 0.5) | 0 (0, 0.3) | 0 (0, 0.3) | 0 (0, 0) | 0 (0, 0) | 0 (0, 0) | 0 (0, 0) |
| **7B** | 0 (0, 0.1) | 0 (0, 0.1) | 0 (0, 1.2) | 0 (0, 0.9) | 0 (0, 0.9) | 0 (0, 0.1) | 0 (0, 0) | 0 (0, 0.1) | 0 (0, 0) |
| **15D[d]** | - | - | 0 (0, 0.1) | 0 (0, 0.1) | 0 (0, 0) | 0 (0, 0) | 0 (0, 0) | 0 (0, 0) | 0 (0, 0.1) |

Abbreviations: CI – confidence interval; AOM – acute otitis media; IPD – invasive pneumococcal disease.
[a] Percent point estimates do not sum to 100% given estimate uncertainty ranges.
[b] Non-bacteremic pneumonia. Bacteremic pneumonia included in IPD.
[c] For adult pneumonia, estimates for 6A are for 6A and 6C combined due to serotype-specific urinary antigen detection assay cross-reactivity for these two serotypes.
[d] 15D has only been identified in invasive pneumococcal disease[20] and so was not considered for pediatric, non-invasive samples.

**Table S8. Estimated percent of pneumococcal infections and number of cases potentially preventable by 21-valent pneumococcal conjugate vaccine (PCV21) in children[a] with risk factors for pneumococcal disease**

| Condition | Assuming no 15B and 15C cross-protection | | Assuming complete 15B and 15C cross-protection | |
|---|---|---|---|---|
| | *% pneumococcal isolates (95% CI)[b]* | *No. cases in thousands (95% CI)[c]* | *% pneumococcal isolates (95% CI)* | *No. cases in thousands (95% CI)[b]* |
| AOM[d] | 77.3 (76.1, 78.4) | 64.7 (41.9, 97.4) | 78.9 (77.8, 79.9) | 66.2 (42.9, 99.6) |
| Sinusitis[d] | 76.7 (75.5, 77.9) | 116.6 (63.7, 195.3) | 78.2 (77.2, 79.3) | 119.2 (65.1, 199.6) |
| Pneumonia (outpatients)[e] | 76.7 (75.5, 77.9) | 19.8 (10.8, 33.6) | 78.2 (77.2, 79.3) | 20.3 (11.1, 34.3) |
| Pneumonia (inpatients)[e] | 76.7 (75.5, 77.9) | 0.5 (0.3, 0.8) | 78.2 (77.2, 79.3) | 0.5 (0.3, 0.8) |
| IPD | 75.3 (72.6, 77.8) | 0.1 (0.1, 0.1) | 79.3 (76.7, 81.6) | 0.1 (0.1, 0.2) |

Abbreviations: CI – confidence interval; AOM – acute otitis media; IPD – invasive pneumococcal disease.

[a] Children aged 5–17 years for all conditions except non-bacteremic pneumonia managed in outpatient settings which includes children 2–17 years to ensure adequate sample size for National Ambulatory Medicare Care Survey projection validity.[21] PCV15 or PCV20 currently recommended for children 5–17 years with health conditions placing them at increased risk of pneumococcal disease.[12,13]

[b] Total percent of pneumococcal isolates not excluding PCV13 serotypes.

[c] Considering US population by age group[22] and assuming disease incidence rates as detailed in **Tables S11–S13**, pneumococcal-attributable disease proportions (**Table S4)**, preventable proportions of disease (excluding PCV13 serotypes), percents of pediatric pneumococcal cases occurring among children with risk-based PCV recommendations (**Table S5**), and PCV effectiveness (**Table S6).**

[d] Percent of cases occurring among children with risk-based indications for PCVs for pneumococcal AOM and sinusitis assumed to be consistent with pneumonia (**Table S5**).

[e] Non-bacteremic pneumonia. Bacteremic pneumonia classified as IPD.

**Table S9. Proportion of pneumococcal infections due to serotypes targeted by pneumococcal conjugate vaccine products assuming complete serotype 15B and 15C cross-protection[a,b] and comparison with primary analysis estimates[c] among children 0–17 years**

| Condition | Age group | PCV20 | | PCV21 | | PCV24[d] | | PCV25 | | PCV31 | |
|---|---|---|---|---|---|---|---|---|---|---|---|
| | | *% of isolates (95% CI)* | *Percent change from primary (95% CI)* [c] | *% of isolates (95% CI)* | *Percent change from primary (95% CI)* [c] | *% of isolates (95% CI)* | *Percent change from primary (95% CI)* [c] | *% of isolates (95% CI)* | *Percent change from primary (95% CI)* [c] | *% of isolates (95% CI)* | *Percent change from primary (95% CI)* [c] |
| AOM | 0–17 years | 42.1 (40.9, 43.4) | 37.3 (33.8, 41.0) | 78.9 (77.8, 79.9) | 2.0 (1.6, 2.5) | 45.0 (43.8, 46.3) | 34.1 (30.9, 37.4) | 54.4 (53.2, 55.7) | 26.6 (24.2, 29.2) | 79.5 (78.4, 80.5) | 16.8 (15.4, 18.3) |
| Sinusitis, pneumonia[e] | 0–17 years | 42.6 (41.4, 43.8) | 36.4 (33.0, 39.9) | 78.2 (77.2, 79.3) | 2.0 (1.6, 2.4) | 45.5 (44.3, 46.8) | 33.3 (30.2, 36.5) | 54.9 (53.7, 56.1) | 26.1 (23.7, 28.5) | 79.6 (78.6, 80.6) | 16.6 (15.2, 18.1) |
| IPD | 0–17 years | 61.9 (58.9, 64.7) | 10.6 (8.1, 13.6) | 79.3 (76.7, 81.6) | 5.2 (3.8, 7.0) | 65.2 (62.4, 68.1) | 10.0 (7.7, 12.8) | 74.2 (71.4, 76.8) | 8.7 (6.7, 11.1) | 90.9 (89.0, 92.6) | 7.0 (5.4, 8.9) |

Abbreviations: PCV – pneumococcal conjugate vaccine; CI – confidence interval; AOM – acute otitis media; IPD – invasive pneumococcal disease.

[a] PCV20 includes serotypes 1, 3, 4, 5, 6A/C, 6B, 7F, 8, 9V, 10A, 11A, 12F, 14, 15B, 18C, 19F, 19A, 22F, 23F, 33F. PCV24 includes serotypes 1, 2, 3, 4, 5, 6A/C, 6B, 7F, 8, 9N, 9V, 10A, 11A, 12F, 14, 15B, 17F, 18C, 19F, 19A, 20B, 22F, 23F, 33F. PCV25 includes serotypes 1, 2, 3, 4, 5, 6B, 6A/C, 7F, 8, 9N, 9V, 10A, 12F, 14, 15A, 15B, 16F, 18C, 19F, 19A, 22F, 23F, 24F, 33F, 35B. PCV31 includes serotypes 1, 2, 3, 4, 5, 6A/C, 6B, 7F, 7C, 8, 9N, 9V, 10A, 11A, 12F, 14, 15A, 15B, 16F, 17F, 18C, 19F, 19A, 20B, 22F, 23F, 23A, 23B, 31, 33F, 35B.

[b] PCV15 not included as it does not target serotypes 15B or 15C. PCV21 not considered as it is intended only for use in adults.

[c] Percent change comparing estimates assuming complete 15B/C cross-protection compared with estimates from primary analysis (**Table 1**) which assumed no 15B/C cross-protection.

[d] Also includes Pn-MAPS24v.

[e] Non-bacteremic pneumonia. Bacteremic pneumonia classified as IPD.

**Table S10. Proportion of pneumococcal infections due to serotypes targeted by pneumococcal conjugate vaccine products assuming complete serotype 15B and 15C cross-protection[a,b] and comparison with primary analysis estimates[c] among adults ≥18 years**

| | | PCV20 | | PCV21 | | PCV24[d] | | PCV25 | | PCV31 | |
|---|---|---|---|---|---|---|---|---|---|---|---|
| **Condition** | **Age group** | *% of isolates (95% CI)* | *Percent change from primary (95% CI)* [c] | *% of isolates (95% CI)* | *Percent change from primary (95% CI)* [c] | *% of isolates (95% CI)* | *Percent change from primary (95% CI)* [c] | *% of isolates (95% CI)* | *Percent change from primary (95% CI)* [c] | *% of isolates (95% CI)* | *Percent change from primary (95% CI)* [c] |
| Pneumonia[e] | | | | | | | | | | | |
| | ≥18 years | 53.6 (48.8, 58.2) | 2.6 (1.2, 5.5) | 71.0 (66.1, 75.2) | 3.6 (1.8, 6.3) | 66.8 (62.3, 71.3) | 2.1 (1.0, 4.4) | 63.3 (58.2, 67.9) | 2.2 (1.0, 4.6) | 88.2 (84.2, 91.2) | 1.6 (0.7, 3.3) |
| | 18–49 years | 47.0 (36.8, 56.9) | 3.6 (0.7, 13.2) | 75.0 (62.9, 82.6) | 2.4 (0.4, 8.2) | 64.7 (54.5, 74.1) | 2.6 (0.5, 9.3) | 62.0 (50.4, 71.3) | 2.7 (0.5, 10.0) | 87.5 (76.1, 93.3) | 1.9 (0.4, 6.8) |
| | 50–64 years | 52.6 (45.1, 59.8) | 0.6 (0.0, 4.2) | 68.9 (60.8, 76.0) | 2.5 (0.5, 6.7) | 67.3 (60.0, 74.4) | 0.5 (0.0, 3.3) | 62.2 (53.5, 69.3) | 0.5 (0.0, 3.5) | 86.4 (78.5, 91.1) | 0.4 (0.0, 2.5) |
| | ≥65 years | 58.1 (49.6, 65.5) | 3.1 (0.9, 8.7) | 69.5 (60.5, 76.3) | 4.9 (1.7, 10.8) | 67.1 (59.3, 74.6) | 2.6 (0.8, 7.5) | 64.3 (55.5, 71.4) | 2.8 (0.8, 7.9) | 89.2 (80.9, 93.7) | 2.0 (0.6, 5.6) |
| IPD | | | | | | | | | | | |
| | ≥18 years | 59.1 (58.2, 59.9) | 1.7 (1.4, 2.0) | 83.6 (82.9, 84.2) | 1.9 (1.6, 2.2) | 69.8 (69.0, 70.5) | 1.5 (1.2, 1.7) | 73.9 (73.2, 74.7) | 1.4 (1.1, 1.6) | 94.9 (94.5, 95.3) | 1.1 (0.9, 1.3) |
| | 18–49 years | 63.0 (61.2, 64.8) | 1.4 (1.0, 2.0) | 80.4 (78.9, 81.8) | 1.7 (1.2, 2.3) | 76.5 (74.9, 78.0) | 1.2 (0.8, 1.7) | 76.2 (74.6, 77.8) | 1.2 (0.8, 1.7) | 95.9 (95.1, 96.6) | 0.9 (0.6, 1.3) |
| | 50–64 years | 60.0 (58.6, 61.4) | 1.3 (1.0, 1.8) | 82.7 (81.5, 83.7) | 1.4 (1.0, 1.8) | 72.5 (71.2, 73.8) | 1.1 (0.8, 1.5) | 74.3 (73.0, 75.6) | 1.1 (0.8, 1.5) | 95.0 (94.3, 95.6) | 0.8 (0.6, 1.1) |
| | ≥65 years | 56.2 (54.9, 57.5) | 2.2 (1.7, 2.8) | 85.9 (84.9, 86.8) | 2.4 (2.0, 2.9) | 63.9 (62.6, 65.2) | 2.0 (1.5, 2.5) | 72.3 (71.1, 73.5) | 1.7 (1.4, 2.2) | 94.2 (93.6, 94.8) | 1.3 (1.0, 1.7) |

Abbreviations: PCV – pneumococcal conjugate vaccine; CI – confidence interval; IPD – invasive pneumococcal disease.

[a] PCV20 includes serotypes 1, 3, 4, 5, 6A/C, 6B, 7F, 8, 9V, 10A, 11A, 12F, 14, 15B/C, 18C, 19F, 19A, 22F, 23F, 33F. PCV21 includes serotypes 3, 6A/C, 7F, 8, 9N, 10A, 11A, 12F, 15A, 15B/C, 16F, 17F, 19A, 20A, 22F, 23A, 23B, 24F, 31, 33F, 35B. PCV24 includes serotypes 1, 2, 3, 4, 5, 6A/C, 6B, 7F, 8, 9N, 9V, 10A, 11A, 12F, 14, 15B/C, 17F, 18C, 19F, 19A, 20B, 22F, 23F, 33F. PCV25 includes serotypes 1, 2, 3, 4, 5, 6B, 6A/C, 7F, 8, 9N, 9V, 10A, 12F, 14, 15A, 15B/C, 16F, 18C, 19F, 19A,

22F, 23F, 24F, 33F, 35B. PCV31 includes serotypes 1, 2, 3, 4, 5, 6A/C, 6B, 7F, 7C, 8, 9N, 9V, 10A, 11A, 12F, 14, 15A, 15B/C, 16F, 17F, 18C, 19F, 19A, 20B, 22F, 23F, 23A, 23B, 31, 33F, 35B.

[b] PCV15 not included as it does not target serotypes 15B or 15C.

[c] Percent change comparing estimates assuming complete 15B/C cross-protection compared with estimates from primary analysis (**Table 2**) which assumed no 15B/C cross-protection. .

[d] Also includes Pn-MAPS24v.

[e] Non-bacteremic pneumonia. Bacteremic pneumonia classified as IPD.

**Table S11. Estimated national burdens of invasive pneumococcal disease in 2019**

| Age category[a] | Estimated national number of cases[b] |
|---|---|
| <2 years | 915 |
| 2–4 years | 515 |
| 5–17 years | 749 |
| 18–49 years | 6,036 |
| 50–64 years | 9,813 |
| ≥65 years | 12,781 |

[a] Where age groups from Active Bacterial Core surveillance (ABCs) do not align with those in this study, we took the population-weighted average.
[b] Estimated as ABCs incidence estimates[23] multiplied by corresponding 2019 bridged-race census estimates.[22] Confidence intervals not estimated consistent with ABCs estimates.

**Table S12. Estimated national burdens of all-cause AOM, pneumonia, and sinusitis visits among outpatients in 2019**

| Condition | Age category | Visit incidence per 1000 person-years (95% CI) | | | Estimated national number in millions (95% CI)[d] |
|---|---|---|---|---|---|
| | | *Physician offices and EDs[a]* | *All other outpatient settings[b]* | *Total[c]* | |
| AOM | | | | | |
| | <2 years | 502 (329, 726) | 127 (111, 142) | 628 (455, 853) | 4.8 (3.5, 6.5) |
| | 2–4 years | 300 (199, 431) | 89 (77, 103) | 390 (288, 521) | 4.7 (3.4, 6.2) |
| | 5–17 years | 53 (32, 80) | 21 (15, 28) | 74 (53, 103) | 4.0 (2.8, 5.5) |
| Pneumonia[e] | | | | | |
| | <2 years | 38 (20, 64) | 12 (7, 17) | 49 (31, 76) | 0.4 (0.2, 0.6) |
| | 2–17 years | 9 (6, 14) | 5 (2, 9) | 15 (10, 21) | 1.0 (0.6, 1.3) |
| | 18–49 years | 7 (4, 11) | 7 (4, 11) | 14 (10, 20) | 2.0 (1.3, 2.7) |
| | 50–64 years | 18 (10, 29) | 29 (23, 36) | 48 (37, 61) | 3.0 (2.4, 3.8) |
| | ≥65 years | 38 (21, 63) | 98 (85, 112) | 137 (115, 164) | 7.4 (6.2 8.9) |
| Sinusitis | | | | | |
| | 5–17 years | 43 (28, 63) | 14 (10, 20) | 58 (41, 78) | 3.1 (2.2, 4.2) |
| | 18–49 years | 49 (38, 61) | 30 (22, 39) | 79 (65, 93) | 10.9 (9.0, 12.9) |
| | 50–64 years | 60 (44, 79) | 31 (23, 39) | 90 (73, 111) | 5.7 (4.6, 7.0) |
| | ≥65 years | 56 (39, 76) | 31 (24, 39) | 87 (69, 109) | 4.7 (3.7, 5.9) |

Abbreviations: ED – emergency department; CI – confidence interval; AOM – acute otitis media.

[a] Estimated using 2016 and 2019 estimates from the National Ambulatory Medical Care Survey and the National Hospital Ambulatory Medical Care Survey (NAMCS/NHAMCS) and 2016 and 2019 (Vintage 2020) bridged race census estimates.

[b] For age categories <65 years, calculated as the weighted average of estimates from 2016–2019 Meritage MarketScan Commercial and Medicaid datasets. Ages ≥65 years are not included in MarketScan Commercial and Medicaid datasets. To estimate visits to all other outpatient settings for adults ≥65 years for each condition, we used the ratio of office and ED visit incidence in NAMCS/NHAMCS to visit incidence rates from all other outpatient settings in the MarketScan Datasets among adults 50–64 years to impute MarketScan visit incidence in adults ≥65 years, taking the weighted average of resulting estimates from the Commercial and Medicaid datasets. Weights were based on the percent of US residents with any private insurance in 2019: 0–17 years of age – 63.6%; 18–64 years of age – 74.4%; ≥65 years of age – 51.6%.[24]

[c] Sum of incidence in physician offices and EDs and all other settings. Sum of composite values table may not equal total incidence value due to rounding and uncertainty propagation methods.

[d] Calculated by multiplying total incidence estimates by 2019 bridged-race census estimates.

[e] Assumed to be only non-bacteremic pneumonia.

**Table S13. Estimated national burdens of all-cause pneumonia among inpatients in 2019**

| Age category | Projected national number of cases (95% CI)[a,b] |
|---|---|
| <2 years | 22,195 (20,255, 24,135) |
| 2–4 years | 21,255 (18,914, 23,596) |
| 5–17 years | 25,835 (23,235, 28,435) |
| 18–49 years | 90,160 (87,829, 92,491) |
| 50–64 years | 159,240 (155,377, 163,103 |
| ≥65 years | 457,565 (447,197, 467,932) |

Abbreviations: CI – confidence interval; N – number.

[a] Estimated from the 2019 Healthcare Cost and Utilization Project National Inpatient Sample.[25] To propagate uncertainty when including multiple inputs in estimates of preventable burdens, we fit the NIS-derived values to beta distributions and sampled 4,900,000 draws from these distributions.

[b] Identified using *International Classification of Diseases, 10th revision, Clinical Modification* codes A01.03, A02.22, A15, A15.0, A15.7, A15.8, A15.9, A20.2, A21.2, A22.1, A37.01, A37.11, A37.81, A37.91, A42.0, A43.0, A48.1, A50.04, A54.84, B0.12, B05.2, B06.81, B2.50, B37.1, B38.0, B38.1, B38.2, B39.0, B39.1, B39.2, B44.0, B44.1, B58.3, B59, B77.81, J09.X1, J10.0, J10.00, J10.01, J10.08, J11.0, J11.00, J11.08, J12, J12.0, J12.1, J12.2, J12.3, J12.8, J12.81, J12.82, J12.89, J12.9, J13, J14, J15, J15.0, J15.1, J15.2, J15.20, J15.21, J15.211, J15.212, J15.29, J15.3, J15..4, J15.5, J15.6, J15.7, J15.8, J15.9, J16, J16.0, J16.8, J17, J18, J18.0, J18.1, J18.2, J18.8, J18.9, J85.1, P23, P23.0, P23.1, P23.2, P23.3, P23.4, P23.5, P23.6, P23.8, P23.9.

**Table S14. Preventable proportion of pneumococcal infections due to serotypes targeted by pneumococcal conjugate vaccines[a,b] excluding PCV13 serotypes + 6C**

| | | Percent of pneumococcal isolates (95% CI) | | | | | |
|---|---|---|---|---|---|---|---|
| **Condition** | **Age group** | *PCV15* | *PCV20* | *PCV21* [c] | *PCV24*[d] | *PCV25* | *PCV31* |
| AOM | | | | | | | |
| | 0–17 years | 4.2 (3.7, 4.7) | 18.8 (17.8, 19.7) | – – | 21.7 (20.7, 22.7) | 31.1 (29.9, 32.2) | 56.1 (54.8, 57.4) |
| Sinusitis, pneumonia[e] | | | | | | | |
| | 0–17 years | 4.2 (3.7, 4.7) | 18.7 (17.7, 19.6) | – – | 21.6 (20.6, 22.6) | 31.0 (29.9, 32.1) | 55.7 (54.4, 56.9) |
| Pneumonia[e] | | | | | | | |
| | ≥18 years | 8.7 (6.2, 11.9) | 18.3 (14.7, 22.5) | 45.8 (41.0, 51.0) | 31.5 (27.1, 36.5) | 28.0 (23.7, 32.7) | 52.9 (47.9, 57.9) |
| | 18–49 years | 8.3 (3.3, 15.3) | 16.5 (10.1, 26.0) | 50.8 (40.1, 62.5) | 34.3 (25.0, 45.3) | 31.5 (22.0, 42.0) | 57.0 (46.3, 67.5) |
| | 50–64 years | 11.1 (6.7, 17.0) | 19.3 (13.8, 26.4) | 45.5 (37.7, 53.9) | 33.9 (27.0, 42.3) | 28.8 (21.9, 36.5) | 52.9 (44.6, 60.8) |
| | ≥65 years | 5.9 (2.5, 10.9) | 17.6 (12.3, 25.2) | 41.4 (33.4, 50.1) | 26.7 (19.9, 35.0) | 23.7 (17.7, 31.2) | 48.7 (40.0, 57.0) |
| IPD | | | | | | | |
| | 0–17 years | 15.6 (13.6, 17.9) | 29.8 (27.0, 32.6) | – – | 33.2 (30.4, 36.1) | 42.1 (39.1, 45.0) | 58.9 (55.9, 61.8) |
| | ≥18 years | 14.2 (13.6, 14.8) | 30.0 (29.2, 30.8) | 59.8 (59.0, 60.7) | 40.7 (39.8, 41.5) | 44.8 (44.0, 45.7) | 65.8 (65.0, 66.6) |
| | 18–49 years | 13.2 (11.9, 14.4) | 32.4 (30.7, 34.2) | 58.9 (57.0, 60.7) | 45.9 (44.0, 47.8) | 45.6 (43.8, 47.5) | 65.3 (63.5, 67.1) |
| | 50–64 years | 13.4 (12.6, 14.2) | 30.6 (29.3, 32.0) | 59.4 (58.0, 60.8) | 43.2 (41.7, 44.6) | 45.0 (43.5, 46.4) | 65.6 (64.3, 67.0) |
| | ≥65 years | 15.3 (14.3, 16.2) | 28.1 (26.9, 29.3) | 60.7 (59.3, 61.9) | 35.8 (34.5, 37.1) | 44.2 (42.9, 45.6) | 66.2 (64.9, 67.4) |

Abbreviations: CI – confidence interval; PCV – pneumococcal conjugate vaccine; AOM – acute otitis media; IPD – invasive pneumococcal disease.

[a] Proportions include all serotypes except those targeted by PCV13 (1, 3, 4, 5, 6A, 6B, 7F, 9V, 14, 18C, 19A, 19F, 23F) and 6C as we assume no additional benefit from these vaccines compared with PCV13. PCV15 includes serotypes 22F, 33F. PCV20 includes serotypes 8, 10A, 11A, 12F, 15B, 22F, 33F. PCV21 includes serotypes 8, 9N, 10A, 11A, 12F, 15A, 15C, 16F, 17F, 20A, 22F, 23A, 23B, 24F, 31, 33F, 35B. PCV24 includes serotypes 2, 6B, 8, 9N, 10A, 11A, 12F, 15B, 17F, 20B, 22F, 33F. PCV25 includes serotypes 2, 9N, 10A, 12F, 15A, 15B, 16F, 22F, 24F, 33F, 35B. PCV31 includes serotypes 2, 7C, 8, 9N, 10A, 11A, 12F, 15A, 15B, 16F, 17F, 20B, 22F, 23A, 23B, 31, 33F, 35B.

[b] Assuming no 15B and 15C cross-protection.

[c] Estimates for PCV21 only presented for adults as PCV21 is not indicated for use in children.

[d] Also includes Pn-MAPS24v.

[e] Non-bacteremic pneumonia. Bacteremic pneumonia classified as IPD.

**Table S15. Preventable proportion of pneumococcal infections due to serotypes targeted by pneumococcal conjugate vaccines[a] excluding PCV13 serotypes + 6C assuming complete serotype 15B and 15C cross-protection**

| | | Percent of pneumococcal isolates (95% CI) | | | | | |
|---|---|---|---|---|---|---|---|
| **Condition** | **Age group** | *PCV15* | *PCV20* | *PCV21[b]* | *PCV24[c]* | *PCV25* | *PCV31* |
| AOM | | | | | | | |
| | 0–17 years | 4.2 (3.7, 4.7) | 30.2 (29.1, 31.3) | – – | 33.1 (32.0, 34.3) | 42.5 (41.3, 43.7) | 67.6 (66.4, 68.7) |
| Sinusitis, pneumonia[d] | | | | | | | |
| | 0–17 years | 4.2 (3.7, 4.7) | 30.0 (28.9, 31.1) | – – | 32.9 (31.8, 34.1) | 42.3 (41.1, 43.5) | 67.0 (65.8, 68.2) |
| Pneumonia[d] | | | | | | | |
| | ≥18 years | 8.7 (6.2, 11.9) | 19.8 (16.0, 24.1) | 48.4 (43.5, 53.6) | 33.0 (28.5, 38.0) | 29.4 (25.1, 34.2) | 54.3 (49.4, 59.3) |
| | 18–49 years | 8.3 (3.3, 15.3) | 18.3 (11.5, 28.8) | 52.9 (41.8, 64.3) | 36.3 (27.0, 47.4) | 33.3 (23.6, 44.2) | 58.9 (48.6, 69.4) |
| | 50–64 years | 11.1 (6.7, 17.0) | 19.7 (14.2, 27.0) | 47.4 (39.2, 55.8) | 34.4 (27.3, 42.8) | 29.2 (22.5, 37.1) | 53.4 (45.2, 61.3) |
| | ≥65 years | 5.9 (2.5, 10.9) | 19.5 (13.7, 27.5) | 45.0 (36.5, 53.5) | 28.7 (21.5, 37.1) | 25.7 (19.2, 33.4) | 50.8 (41.7, 58.9) |
| IPD | | | | | | | |
| | 0–17 years | 15.6 (13.6, 17.9) | 35.8 (32.8, 38.6) | – – | 39.1 (36.2, 42.1) | 48.1 (45.0, 51.0) | 64.8 (61.9, 67.6) |
| | ≥18 years | 14.2 (13.6, 14.8) | 31.0 (30.2, 31.8) | 61.4 (60.5, 62.2) | 41.7 (40.8, 42.5) | 45.8 (45.0, 46.7) | 66.8 (66.0, 67.6) |
| | 18–49 years | 13.2 (11.9, 14.4) | 33.3 (31.6, 35.1) | 60.2 (58.4, 62.1) | 46.8 (44.9, 48.6) | 46.5 (44.7, 48.4) | 66.2 (64.4, 68.0) |
| | 50–64 years | 13.4 (12.6, 14.2) | 31.4 (30.1, 32.8) | 60.5 (59.1, 61.9) | 44.0 (42.5, 45.4) | 45.8 (44.3, 47.2) | 66.4 (65.1, 67.8) |
| | ≥65 years | 15.3 (14.3, 16.2) | 29.3 (28.1, 30.6) | 62.7 (61.4, 63.9) | 37.0 (35.8, 38.3) | 45.5 (44.1, 46.8) | 67.4 (66.1, 68.6) |

Abbreviations: PCV – pneumococcal conjugate vaccine; CI – confidence interval; AOM – acute otitis media; IPD – invasive pneumococcal disease.

[a] Proportions include all serotypes except those targeted by PCV13 (1, 3, 4, 5, 6A, 6B, 7F, 9V, 14, 18C, 19A, 19F, 23F) and 6C as we assume no additional benefit from these vaccines compared with PCV13. PCV15 includes serotypes 22F, 33F. PCV20 includes serotypes 8, 10A, 11A, 12F, 15B, 22F, 33F. PCV21 includes serotypes 8, 9N, 10A, 11A, 12F, 15A, 15C, 16F, 17F, 20A, 22F, 23A, 23B, 24F, 31, 33F, 35B. PCV24 includes serotypes 2, 6B, 8, 9N, 10A, 11A, 12F, 15B, 17F, 20B, 22F, 33F. PCV25 includes serotypes 2, 9N, 10A, 12F, 15A, 15B, 16F, 22F, 24F, 33F, 35B. PCV31 includes serotypes 2, 7C, 8, 9N, 10A, 11A, 12F, 15A, 15B, 16F, 17F, 20B, 22F, 23A, 23B, 31, 33F, 35B.

[b] Estimates for PCV21 only presented for adults as PCV21 is not indicated for use in children.
[c] Also includes Pn-MAPS24v.
[d] Non-bacteremic pneumonia. Bacteremic pneumonia classified as IPD.

**Table S16. Estimated number of cases potentially preventable by pneumococcal conjugate vaccines[a,b] in children 0–17 years**

| Condition | Age category | Number of cases in thousands (95% CI)[c] | | | | |
|---|---|---|---|---|---|---|
| | | *PCV15* | *PCV20* | *PCV24*[d] | *PCV25* | *PCV31* |
| AOM | | | | | | |
| | <2 years[e] | 21.8 (14.2, 32.7) | 98.4 (64.9, 145.7) | 113.7 (75.0, 168.2) | 163.0 (107.7, 240.8) | 294.4 (194.7, 434.6) |
| | 2–4 years[f] | 21.3 (14.1, 31.5) | 96.1 (64.3, 140.4) | 111.0 (74.3, 162.1) | 159.2 (106.7, 232.1) | 287.6 (192.9, 418.8) |
| | 5–17 years, all[g] | 18.1 (11.6, 27.6) | 81.8 (53.1, 122.8) | 94.5 (61.4, 141.8) | 135.4 (88.0, 203.0) | 244.6 (159.2, 366.4) |
| | 5–17 years, w/ risk conditions[h] | 3.9 (2.5, 6.0) | 17.7 (11.4, 26.7) | 20.4 (13.2, 30.8) | 29.2 (18.9, 44.1) | 52.8 (34.2, 79.5) |
| | Total, all[i,j] | 61.7 (43.9, 84.8) | 278.8 (201.5, 375.9) | 322.0 (232.9, 434.0) | 461.6 (334.3, 621.2) | 833.9 (604.7, 1,120.4) |
| | Total, PCVs recommended[i,k] | 47.3 (33.3, 65.7) | 213.8 (152.9, 291.8) | 246.9 (176.7, 336.8) | 354.0 (253.6, 482.2) | 639.4 (458.7, 869.8) |
| Sinusitis[l] | | | | | | |
| | 5–17 years, all[g] | 32.9 (17.8, 55.5) | 147.6 (80.7, 246.8) | 170.7 (93.4, 285.3) | 245.1 (134.2, 409.1) | 440.5 (241.2, 734.6) |
| | 5–17 years, w/ risk conditions[h] | 7.1 (3.8, 12.0) | 31.9 (17.4, 53.5) | 36.9 (20.1, 61.8) | 52.9 (28.9, 88.7) | 95.1 (51.9, 159.3) |
| Pneumonia (outpatients)[m] | | | | | | |
| | <2 years[d] | 2.2 (1.1, 4.0) | 9.9 (5.2, 17.8) | 11.4 (6.0, 20.6) | 16.4 (8.6, 29.5) | 29.5 (15.4, 53.0) |
| | 2–17 years, all[f] | 5.6 (3.0, 9.5) | 25.1 (13.7, 42.4) | 29.0 (15.9, 49.0) | 41.6 (22.8, 70.3) | 74.8 (41.0, 126.2) |
| | 2–17 years, w/ risk conditions[n] | 1.2 (0.7, 2.1) | 5.4 (3.0, 9.2) | 6.3 (3.4, 10.6) | 9.0 (4.9, 15.2) | 16.2 (8.8, 27.4) |
| | Total, all[h,i] | 7.9 (4.5, 12.8) | 35.3 (20.4, 57.0) | 40.8 (23.5, 65.8) | 58.6 (33.8, 94.4) | 105.2 (60.8, 169.5) |
| | Total, PCVs recommended[h,l] | 3.4 (1.9, 5.7) | 15.4 (8.8, 25.4) | 17.9 (10.2, 29.4) | 25.6 (14.7, 42.2) | 46.1 (26.4, 75.7) |
| Pneumonia (inpatients)[m] | | | | | | |
| | <2 years[d] | 0.1 (0.1, 0.2) | 0.5 (0.3, 0.8) | 0.6 (0.4, 0.9) | 0.9 (0.5, 1.3) | 1.6 (0.9, 2.4) |
| | 2–4 years[e] | 0.1 (0.1, 0.2) | 0.5 (0.3, 0.8) | 0.6 (0.4, 0.9) | 0.9 (0.5, 1.3) | 1.6 (0.9, 2.4) |

| Condition | Age category | Number of cases in thousands (95% CI)[c] | | | | |
|---|---|---|---|---|---|---|
| | | *PCV15* | *PCV20* | *PCV24*[d] | *PCV25* | *PCV31* |
| | 5–17 years, all[f] | 0.1 (0.1, 0.2) | 0.6 (0.4, 1.0) | 0.7 (0.4, 1.1) | 1.0 (0.6, 1.6) | 1.9 (1.1, 2.8) |
| | 5–17 years, w/ risk conditions[g] | 0.0 (0.0, 0.0) | 0.1 (0.1, 0.2) | 0.2 (0.1, 0.2) | 0.2 (0.1, 0.3) | 0.4 (0.2, 0.6) |
| | Total, all[h,i] | 0.4 (0.2, 0.6) | 1.7 (1.0, 2.5) | 1.9 (1.2, 2.9) | 2.8 (1.7, 4.2) | 5.0 (3.0, 7.5) |
| | Total, PCVs recommended[h,j] | 0.3 (0.2, 0.4) | 1.2 (0.7, 1.8) | 1.4 (0.8, 2.1) | 2.0 (1.2, 3.0) | 3.5 (2.1, 5.3) |
| IPD | | | | | | |
| | <2 years[d] | 0.1 (0.1, 0.2) | 0.2 (0.2, 0.3) | 0.3 (0.2, 0.3) | 0.3 (0.3, 0.4) | 0.5 (0.4, 0.5) |
| | 2–4 years[e] | 0.1 (0.1, 0.1) | 0.1 (0.1, 0.2) | 0.2 (0.1, 0.2) | 0.2 (0.2, 0.2) | 0.3 (0.2, 0.3) |
| | 5–17 years, all[f] | 0.1 (0.1, 0.1) | 0.2 (0.2, 0.2) | 0.2 (0.2, 0.3) | 0.3 (0.2, 0.3) | 0.4 (0.3, 0.4) |
| | 5–17 years, w/ risk conditions[g] | 0.0 (0.0, 0.0) | 0.1 (0.0, 0.1) | 0.1 (0.1, 0.1) | 0.1 (0.1, 0.1) | 0.1 (0.1, 0.1) |
| | Total, all[h,i] | 0.3 (0.3, 0.4) | 0.6 (0.5, 0.7) | 0.7 (0.6, 0.7) | 0.8 (0.7, 0.9) | 1.2 (1.0, 1.3) |
| | Total, PCVs recommended[h,j] | 0.2 (0.2, 0.3) | 0.4 (0.4, 0.5) | 0.5 (0.4, 0.6) | 0.6 (0.5, 0.7) | 0.9 (0.8, 1.0) |

Abbreviations: CI – confidence interval; PCV – pneumococcal conjugate vaccine; AOM – acute otitis media; IPD – invasive pneumococcal disease.

[a] PCV15 includes serotypes 1, 3, 4, 5, 6A/C, 6B, 7F, 9V, 14, 18C, 19F, 19A, 22F, 23F, 33F. PCV20 includes serotypes 1, 3, 4, 5, 6A/C, 6B, 7F, 8, 9V, 10A, 11A, 12F, 14, 15B, 18C, 19F, 19A, 22F, 23F, 33F. PCV24 includes serotypes 1, 2, 3, 4, 5, 6A/C, 6B, 7F, 8, 9N, 9V, 10A, 11A, 12F, 14, 15B, 17F, 18C, 19F, 19A, 20B, 22F, 23F, 33F. PCV25 includes serotypes 1, 2, 3, 4, 5, 6B, 6A/C, 7F, 8, 9N, 9V, 10A, 12F, 14, 15A, 15B, 16F, 18C, 19F, 19A, 22F, 23F, 24F, 33F, 35B. PCV31 includes serotypes 1, 2, 3, 4, 5, 6A/C, 6B, 7F, 7C, 8, 9N, 9V, 10A, 11A, 12F, 14, 15A, 15B, 16F, 17F, 18C, 19F, 19A, 20B, 22F, 23F, 23A, 23B, 31, 33F, 35B.

[b] Estimates for PCV21 not presented as PCV21 is not indicated for use in children.

[c] Considering US population by age group[22] and assuming disease incidence rates as detailed in **Tables S11–S13**, pneumococcal-attributable disease proportions as detailed in **Table S4**, PCV preventable proportion of pneumococci by condition and age-group as detailed in **Table S14**, PCV effectiveness as detailed in **Table S6,** and no serotype 15B and 15C cross-protection.

[d] Also includes Pn-MAPS24v.

[e] Vaccination with PCV15 or PCV20 currently recommended for all children 2–23 months without contraindications for vaccination.[12,13]

[f] Vaccination with PCV15 or PCV20 currently recommended for children 24–59 months with incomplete PCV vaccination status.[12,13]

[g] All children aged 5–17 years.

[h] Children 5–17 years with underlying health conditions placing them at increased risk of pneumococcal disease. Proportions of children with at risk conditions derived from Pelton et al. 2014[10] and detailed in **Table S5**. Vaccination with PCV15 or PCV20 recommended for children 5–17 years with health conditions placing them at increased risk of pneumococcal disease.[12,13]

[i] Component values may not sum to total due to rounding.

[j] Total for all children.

[k] Total for all children aged <5 years and children 5–17 years with at-risk conditions for AOM, inpatient pneumonia, and IPD. Total for all children aged <2 years and children 2–17 years with at-risk conditions for outpatient pneumonia.

[l] Sinusitis not considered in children aged <5 years as estimates from NAMCS/NHAMCS did not meet sufficient sample size criteria for validity among children <5 years.[21]

[m] Non-bacteremic pneumonia. Bacteremic pneumonia classified as IPD.

[n] NAMCS/NHAMCS sample sizes for children 2–4 years and 5–17 years were insufficient to meet criteria for validity;[21] we considered these age groups together to ensure adequate sample size. The burden of children 2–17 years with risk conditions was estimated as the proportion of children 5–17 years with risk conditions from Pelton et al. 2014[10] (**Table S5**) multiplied by the total burden at risk in this age group.

**Table S17. Estimated number of cases potentially preventable by pneumococcal conjugate vaccines[a] assuming complete serotype 15B and 15C cross-protection in children 0–17 years**

| | | Number of cases in thousands (95% CI)[c] | | | | |
|---|---|---|---|---|---|---|
| **Condition** | **Age category** | *PCV15* | *PCV20* | *PCV24*[d] | *PCV25* | *PCV31* |
| AOM | | | | | | |
| | <2 years[e] | 21.8 (14.2, 32.7) | 158.5 (104.7, 234.2) | 173.8 (114.8, 256.8) | 223.1 (147.5, 329.4) | 354.5 (234.5, 523.1) |
| | 2–4 years[f] | 21.3 (14.1, 31.5) | 154.8 (103.7, 225.8) | 169.7 (113.7, 247.5) | 217.9 (146.1, 317.4) | 346.3 (232.3, 504.1) |
| | 5–17 years, all[g] | 18.1 (11.6, 27.6) | 131.7 (85.6, 197.5) | 144.4 (93.9, 216.5) | 185.3 (120.6, 277.7) | 294.6 (191.7, 441.1) |
| | 5–17 years, w/ risk conditions[h] | 3.9 (2.5, 6.0) | 28.4 (18.4, 42.9) | 31.2 (20.2, 47.0) | 40.0 (25.9, 60.3) | 63.6 (41.2, 95.7) |
| | Total, all[i,j] | 61.7 (43.9, 84.8) | 448.9 (325.1, 604.2) | 492.2 (356.5, 662.2) | 631.8 (457.9, 849.4) | 1,004.1 (728.3, 1,348.5) |
| | Total, PCVs recommended[i,k] | 47.3 (33.3, 65.7) | 344.2 (246.7, 469.0) | 377.4 (270.5, 514.0) | 484.5 (347.4, 659.3) | 770.0 (552.5, 1,047.1) |
| Sinusitis[l] | | | | | | |
| | 5–17 years, all[g] | 32.9 (17.8, 55.5) | 237.6 (130.0, 396.6) | 260.6 (142.7, 435.1) | 335.0 (183.4, 558.9) | 530.4 (290.5, 884.3) |
| | 5–17 years, w/ risk conditions[h] | 7.1 (3.8, 12.0) | 51.3 (28.0, 86.0) | 56.3 (30.7, 94.3) | 72.3 (39.5, 121.2) | 114.5 (62.5, 191.8) |
| Pneumonia (outpatients)[m] | | | | | | |
| | <2 years[d] | 2.2 (1.1, 4.0) | 15.9 (8.3, 28.6) | 17.4 (9.1, 31.4) | 22.4 (11.7, 40.3) | 35.5 (18.5, 63.8) |
| | 2–17 years, all[f] | 5.6 (3.0, 9.5) | 40.4 (22.1, 68.1) | 44.3 (24.3, 74.8) | 56.9 (31.2, 96.0) | 90.1 (49.4, 152.0) |
| | 2–17 years, w/ risk conditions[n] | 1.2 (0.7, 2.1) | 8.7 (4.8, 14.8) | 9.6 (5.2, 16.2) | 12.3 (6.7, 20.8) | 19.5 (10.6, 33.0) |
| | Total, all[h,i] | 7.9 (4.5, 12.8) | 56.8 (32.8, 91.5) | 62.3 (36.0, 100.4) | 80.0 (46.3, 129.0) | 126.7 (73.3, 204.1) |
| | Total, PCVs recommended[h,l] | 3.4 (1.9, 5.7) | 24.8 (14.2, 40.9) | 27.3 (15.6, 44.9) | 35.0 (20.0, 57.6) | 55.5 (31.7, 91.2) |
| Pneumonia (inpatients)[m] | | | | | | |
| | <2 years[d] | 0.1 (0.1, 0.2) | 0.8 (0.5, 1.3) | 0.9 (0.5, 1.4) | 1.2 (0.7, 1.8) | 1.9 (1.1, 2.9) |
| | 2–4 years[e] | 0.1 (0.1, 0.2) | 0.9 (0.6, 1.3) | 0.9 (0.6, 1.4) | 1.2 (0.7, 1.8) | 1.9 (1.1, 2.9) |
| | 5–17 years, all[f] | 0.1 (0.1, 0.2) | 1.0 (0.6, 1.5) | 1.1 (0.7, 1.7) | 1.4 (0.9, 2.2) | 2.3 (1.4, 3.4) |

| Condition | Age category | Number of cases in thousands (95% CI)[c] | | | | |
|---|---|---|---|---|---|---|
| | | *PCV15* | *PCV20* | *PCV24*[d] | *PCV25* | *PCV31* |
| | 5–17 years, w/ risk conditions[g] | 0.0 (0.0, 0.0) | 0.2 (0.1, 0.3) | 0.2 (0.1, 0.4) | 0.3 (0.2, 0.5) | 0.5 (0.3, 0.7) |
| | Total, all[h,i] | 0.4 (0.2, 0.6) | 2.7 (1.6, 4.1) | 3.0 (1.8, 4.5) | 3.8 (2.3, 5.7) | 6.1 (3.6, 9.1) |
| | Total, PCVs recommended[h,j] | 0.3 (0.2, 0.4) | 1.9 (1.1, 2.9) | 2.1 (1.2, 3.1) | 2.7 (1.6, 4.0) | 4.3 (2.5, 6.4) |
| IPD | | | | | | |
| | <2 years[d] | 0.1 (0.1, 0.2) | 0.3 (0.2, 0.3) | 0.3 (0.3, 0.4) | 0.4 (0.3, 0.4) | 0.5 (0.5, 0.6) |
| | 2–4 years[e] | 0.1 (0.1, 0.1) | 0.2 (0.1, 0.2) | 0.2 (0.2, 0.2) | 0.2 (0.2, 0.2) | 0.3 (0.3, 0.3) |
| | 5–17 years, all[f] | 0.1 (0.1, 0.1) | 0.2 (0.2, 0.3) | 0.3 (0.2, 0.3) | 0.3 (0.3, 0.4) | 0.4 (0.4, 0.5) |
| | 5–17 years, w/ risk conditions[g] | 0.0 (0.0, 0.0) | 0.1 (0.1, 0.1) | 0.1 (0.1, 0.1) | 0.1 (0.1, 0.1) | 0.1 (0.1, 0.2) |
| | Total, all[h,i] | 0.3 (0.3, 0.4) | 0.7 (0.6, 0.8) | 0.8 (0.7, 0.9) | 1.0 (0.8, 1.1) | 1.3 (1.1, 1.4) |
| | Total, PCVs recommended[h,j] | 0.2 (0.2, 0.3) | 0.5 (0.5, 0.6) | 0.6 (0.5, 0.7) | 0.7 (0.6, 0.8) | 1.0 (0.8, 1.1) |

Abbreviations: CI – confidence interval; PCV – pneumococcal conjugate vaccine; AOM – acute otitis media; IPD – invasive pneumococcal disease.

[a] PCV15 includes serotypes 1, 3, 4, 5, 6A/C, 6B, 7F, 9V, 14, 18C, 19F, 19A, 22F, 23F, 33F. PCV20 includes serotypes 1, 3, 4, 5, 6A/C, 6B, 7F, 8, 9V, 10A, 11A, 12F, 14, 15B/C, 18C, 19F, 19A, 22F, 23F, 33F. PCV24 includes serotypes 1, 2, 3, 4, 5, 6A/C, 6B, 7F, 8, 9N, 9V, 10A, 11A, 12F, 14, 15B/C, 17F, 18C, 19F, 19A, 20B, 22F, 23F, 33F. PCV25 includes serotypes 1, 2, 3, 4, 5, 6B, 6A/C, 7F, 8, 9N, 9V, 10A, 12F, 14, 15A, 15B/C, 16F, 18C, 19F, 19A, 22F, 23F, 24F, 33F, 35B. PCV31 includes serotypes 1, 2, 3, 4, 5, 6A/C, 6B, 7F, 7C, 8, 9N, 9V, 10A, 11A, 12F, 14, 15A, 15B/C, 16F, 17F, 18C, 19F, 19A, 20B, 22F, 23F, 23A, 23B, 31, 33F, 35B.

[b] Estimates for PCV21 not presented as PCV21 is not indicated for use in children.

[c] Considering US population by age group[22] and assuming disease incidence rates as detailed in **Tables S11–S13**, pneumococcal-attributable disease proportions as detailed in **Table S4**, PCV preventable proportion of pneumococci by condition and age-group as detailed in **Table S15**, PCV effectiveness as detailed in **Table S6,** and complete 15B and 15C cross-protection.

[d] Also includes Pn-MAPS24v.

[e] Vaccination with PCV15 or PCV20 currently recommended for all children 2–23 months without contraindications for vaccination.[12,13]

[f] Vaccination with PCV15 or PCV20 currently recommended for children 24–59 months with incomplete PCV vaccination status.[12,13]

[g] All children aged 5–17 years.

[h] Children 5–17 years with underlying health conditions placing them at increased risk of pneumococcal disease. Proportions of children with at risk conditions derived from Pelton et al. 2014[10] and detailed in **Table S5**. Vaccination with PCV15 or PCV20 recommended for children 5–17 years with health conditions placing them at increased risk of pneumococcal disease.[12,13]

[i] Component values may not sum to total due to rounding.

[j] Total for all children.

[k] Total for all children aged <5 years and children 5–17 years with at-risk conditions for AOM, inpatient pneumonia, and IPD. Total for all children aged <2 years and children 2–17 years with at-risk conditions for outpatient pneumonia.

[l] Sinusitis not considered in children aged <5 years as estimates from NAMCS/NHAMCS did not meet sufficient sample size criteria for validity among children <5 years.[21]

[m] Non-bacteremic pneumonia. Bacteremic pneumonia classified as IPD.

[n] NAMCS/NHAMCS sample sizes for children 2–4 years and 5–17 years were insufficient to meet criteria for validity;[21] we considered these age groups together to ensure adequate sample size. The burden of children 2–17 years with risk conditions was estimated as the proportion of children 5–17 years with risk conditions from Pelton et al. 2014[10] (**Table S5**) multiplied by the total burden at risk in this age group.

**Table S18. Estimated number of cases potentially preventable by pneumococcal conjugate vaccines[a] in adults ≥18 years**

| Condition | Age category | Number of cases in thousands (95% CI)[b] | | | | | |
|---|---|---|---|---|---|---|---|
| | | *PCV15* | *PCV20* | *PCV21* | *PCV24*[c] | *PCV25* | *PCV31* |
| Sinusitis | | | | | | | |
| | 18–49 years, all[d] | 116.3 (66.8, 182.6) | 522.4 (303.0, 809.4) | 1,911.2 (1,110.3, 2,952.0) | 604.1 (350.4, 935.5) | 867.4 (503.6, 1,341.3) | 1,558.6 (905.6, 2,408.3) |
| | 18–49 years w/ risk conditions[e] | 50.9 (29.3, 80.0) | 228.7 (132.6, 354.5) | 836.7 (486.0, 1,292.9) | 264.5 (153.4, 409.7) | 379.7 (220.4, 587.5) | 682.4 (396.4, 1,054.6) |
| | 50–64 years, all[d] | 60.8 (34.5, 96.9) | 273.1 (156.6, 429.9) | 998.9 (573.7, 1,568.2) | 315.7 (181.1, 496.9) | 453.4 (260.2, 712.6) | 814.7 (467.8, 1,279.3) |
| | ≥65 years[d,f] | 50.2 (28.3, 80.8) | 225.6 (128.5, 358.7) | 825.2 (470.8, 1,308.7) | 260.8 (148.6, 414.6) | 374.5 (213.5, 594.7) | 673.0 (384.0, 1,067.7) |
| | Total, all[g,h] | 228.3 (133.2, 350.9) | 1,025.9 (603.8, 1,554.2) | 3,753.0 (2,213.0, 5,668.3) | 1,186.4 (698.5, 1,796.2) | 1,703.3 (1,003.6, 2,575.8) | 3,060.9 (1,804.7, 4,624.0) |
| | Total, PCVs recommended[g,i] | 162.7 (94.9, 250.2) | 731.2 (430.1, 1,108.6) | 2,675.0 (1,576.4, 4,043.1) | 845.6 (497.4, 1,281.4) | 1,214.0 (714.9, 1,837.6) | 2,181.8 (1,285.7, 3,298.8) |
| Pneumonia (outpatients)[j] | | | | | | | |
| | 18–49 years, all[d] | 6.8 (2.0, 17.4) | 13.8 (5.2, 31.2) | 42.9 (17.6, 84.0) | 29.0 (11.7, 58.6) | 26.4 (10.5, 53.9) | 48.2 (19.9, 93.6) |
| | 18–49 years w/ risk conditions[e] | 3.0 (0.9, 7.6) | 6.1 (2.3, 13.6) | 18.8 (7.7, 36.8) | 12.7 (5.1, 25.6) | 11.6 (4.6, 23.6) | 21.1 (8.7, 41.0) |
| | 50–64 years, all[d] | 21.4 (8.4, 44.3) | 37.5 (15.8, 72.3) | 88.8 (38.7, 159.7) | 66.3 (28.6, 121.7) | 56.0 (24.0, 103.7) | 103.3 (45.2, 184.3) |
| | ≥65 years[d,f] | 21.4 (6.9, 50.6) | 65.1 (27.4, 126.0) | 153.8 (67.5, 273.0) | 99.2 (42.7, 182.8) | 87.8 (37.7, 162.9) | 180.7 (79.6, 317.3) |
| | Total, all[g,h] | 50.0 (18.4, 108.3) | 117.7 (50.4, 220.7) | 288.8 (128.9, 493.5) | 196.7 (86.6, 347.3) | 172.2 (75.5, 306.0) | 335.8 (150.6, 568.7) |
| | Total, PCVs recommended[g,i] | 46.1 (17.1, 99.3) | 109.7 (47.0, 205.2) | 263.9 (117.7, 451.8) | 179.9 (79.2, 318.1) | 156.9 (68.8, 279.1) | 307.9 (138.0, 522.4) |
| Pneumonia (inpatients)[j] | | | | | | | |
| | 18–49 years, all[d] | 0.2 (0.0, 0.4) | 0.3 (0.1, 0.7) | 1.0 (0.4, 2.0) | 0.7 (0.3, 1.4) | 0.6 (0.2, 1.3) | 1.1 (0.5, 2.2) |
| | 18–49 years w/ risk conditions[e] | 0.1 (0.0, 0.2) | 0.2 (0.1, 0.4) | 0.6 (0.2, 1.1) | 0.4 (0.2, 0.8) | 0.4 (0.1, 0.7) | 0.7 (0.3, 1.3) |

| Condition | Age category | Number of cases in thousands (95% CI)[b] | | | | | |
|---|---|---|---|---|---|---|---|
| | | *PCV15* | *PCV20* | *PCV21* | *PCV24*[c] | *PCV25* | *PCV31* |
| IPD | 50–64 years, all[d] | 0.8 (0.3, 1.6) | 1.4 (0.6, 2.6) | 3.3 (1.5, 5.7) | 2.4 (1.1, 4.3) | 2.1 (0.9, 3.7) | 3.8 (1.7, 6.6) |
| | ≥65 years[d,f] | 1.1 (0.4, 2.5) | 3.3 (1.4, 6.3) | 7.8 (3.5, 13.6) | 5.0 (2.2, 9.1) | 4.5 (1.9, 8.1) | 9.2 (4.1, 15.8) |
| | Total, all[g,h] | 2.0 (0.7, 4.4) | 5.1 (2.2, 9.4) | 12.2 (5.5, 20.7) | 8.2 (3.6, 14.4) | 7.2 (3.2, 12.7) | 14.2 (6.4, 23.9) |
| | Total, PCVs recommended[g,i] | 2.0 (0.7, 4.3) | 4.9 (2.1, 9.1) | 11.8 (5.3, 20.0) | 7.9 (3.5, 13.9) | 6.9 (3.1, 12.3) | 13.8 (6.2, 23.1) |
| | 18–49 years, all[d] | 0.6 (0.4, 0.8) | 1.5 (0.9, 1.9) | 2.7 (1.7, 3.4) | 2.1 (1.3, 2.6) | 2.1 (1.3, 2.6) | 3.0 (1.8, 3.7) |
| | 18–49 years w/ risk conditions[e] | 0.4 (0.3, 0.5) | 1.0 (0.6, 1.3) | 1.9 (1.1, 2.3) | 1.4 (0.9, 1.8) | 1.4 (0.9, 1.8) | 2.1 (1.3, 2.6) |
| | 50–64 years, all[d] | 1.0 (0.6, 1.3) | 2.3 (1.4, 2.9) | 4.5 (2.7, 5.5) | 3.2 (2.0, 4.0) | 3.4 (2.1, 4.2) | 4.9 (3.0, 6.1) |
| | ≥65 years[d,f] | 1.5 (0.9, 1.9) | 2.7 (1.7, 3.4) | 5.9 (3.6, 7.3) | 3.5 (2.1, 4.3) | 4.3 (2.6, 5.4) | 6.5 (4.0, 8.0) |
| | Total, all[g,h] | 3.1 (1.9, 3.9) | 6.5 (4.0, 8.1) | 13.1 (8.0, 16.2) | 8.9 (5.4, 11.0) | 9.8 (6.0, 12.2) | 14.4 (8.8, 17.8) |
| | Total, PCVs recommended[g,i] | 2.9 (1.8, 3.6) | 6.1 (3.7, 7.6) | 12.3 (7.5, 15.2) | 8.2 (5, 10.2) | 9.1 (5.6, 11.3) | 13.5 (8.2, 16.7) |

Abbreviations: CI – confidence interval; PCV – pneumococcal conjugate vaccine; IPD – invasive pneumococcal disease.

[a] PCV15 includes serotypes 1, 3, 4, 5, 6A/C, 6B, 7F, 9V, 14, 18C, 19F, 19A, 22F, 23F, 33F. PCV20 includes serotypes 1, 3, 4, 5, 6A/C, 6B, 7F, 8, 9V, 10A, 11A, 12F, 14, 15B, 18C, 19F, 19A, 22F, 23F, 33F. PCV21 includes serotypes 3, 6A/C, 7F, 8, 9N, 10A, 11A, 12F, 15A, 15C, 16F, 17F, 19A, 20A, 22F, 23A, 23B, 24F, 31, 33F, 35B. PCV24 includes serotypes 1, 2, 3, 4, 5, 6A/C, 6B, 7F, 8, 9N, 9V, 10A, 11A, 12F, 14, 15B, 17F, 18C, 19F, 19A, 20B, 22F, 23F, 33F. PCV25 includes serotypes 1, 2, 3, 4, 5, 6B, 6A/C, 7F, 8, 9N, 9V, 10A, 12F, 14, 15A, 15B, 16F, 18C, 19F, 19A, 22F, 23F, 24F, 33F, 35B. PCV31 includes serotypes 1, 2, 3, 4, 5, 6A/C, 6B, 7F, 7C, 8, 9N, 9V, 10A, 11A, 12F, 14, 15A, 15B, 16F, 17F, 18C, 19F, 19A, 20B, 22F, 23F, 23A, 23B, 31, 33F, 35B.

[b] Considering US population by age group[22] and assuming disease incidence rates as detailed in **Tables S11–S13**, pneumococcal-attributable disease proportions as detailed in **Table S4**, PCV preventable proportion of pneumococci by condition and age-group as detailed in **Table S14**, PCV effectiveness as detailed in **Table S6,** and no serotype 15B and 15C cross-protection.

[c] Also includes Pn-MAPS24v.

[d] All adults within age group.

[e] Adults within age group with underlying health conditions placing them at increased risk of pneumococcal disease. Proportions of adults with at risk conditions derived from Grant et al. 2023[11] and detailed in **Table S5**. Vaccination with PCV15 (in addition to 23-valent pneumococcal polysaccharide vaccine [PPSV23]), PCV20, or PCV21 recommended for adults 18–49 years with health conditions placing them at increased risk of pneumococcal disease.[14,15]

[f] Vaccination with PCV15 (plus PPSV23), PCV20, or PCV21 recommended for all adults ≥65 years without contraindications for vaccination.[14,15]

[g] Component values may not sum to total due to rounding.

[h] Total for all adults.

[i] Total for adults 18–49 years with at-risk conditions and all adults ≥50 years.

[j] Non-bacteremic pneumonia. Bacteremic pneumonia classified as IPD.

**Table S19. Estimated number of cases potentially preventable by pneumococcal conjugate vaccines[a] assuming complete serotype 15B and 15C cross-protection in adults ≥18 years**

| Condition | Age category | Number of cases in thousands (95% CI)[b] | | | | | |
|---|---|---|---|---|---|---|---|
| | | *PCV15* | *PCV20* | *PCV21* | *PCV24*[c] | *PCV25* | *PCV31* |
| Sinusitis | | | | | | | |
| | 18–49 years, all[d] | 116.3 (66.8, 182.6) | 840.7 (487.9, 1300.3) | 1,953.6 (1,135.0, 3,017.3) | 922.4 (535.3, 1,426.4) | 1,185.7 (688.6, 1,832.4) | 1,876.9 (1,090.4, 2,899.1) |
| | 18–49 years w/ risk conditions[e] | 50.9 (29.3, 80.0) | 368.1 (213.5, 569.5) | 855.3 (496.8, 1,321.5) | 403.8 (234.3, 624.7) | 519.1 (301.3, 802.5) | 821.7 (477.3, 1,269.7) |
| | 50–64 years, all[d] | 60.8 (34.5, 96.9) | 439.4 (252.2, 690.8) | 1021.1 (586.5, 1,603.0) | 482.1 (276.7, 757.8) | 619.7 (355.8, 973.6) | 981.0 (563.5, 1,540.2) |
| | ≥65 years[d,f] | 50.2 (28.3, 80.8) | 363.0 (206.9, 576.4) | 843.5 (481.3, 1,337.8) | 398.3 (227.0, 632.3) | 511.9 (291.9, 812.4) | 810.4 (462.4, 1,285.3) |
| | Total, all[g,h] | 228.3 (133.2, 350.9) | 1,651.0 (972.4, 2,497.0) | 3,836.5 (2,262.3, 5,793.5) | 1,811.4 (1,067.1, 2,739.1) | 2,328.3 (1,372.2, 3,518.4) | 3,686.1 (2,173.5, 5,566.6) |
| | Total, PCVs recommended[g,i] | 162.7 (94.9, 250.2) | 1,176.7 (692.8, 1,781.1) | 2,734.5 (1,611.5, 4,132.6) | 1,291.0 (760.1, 1,953.7) | 1,659.5 (977.7, 2,509.6) | 2,627.2 (1,548.3, 3,970.8) |
| Pneumonia (outpatients)[j] | | | | | | | |
| | 18–49 years, all[d] | 6.8 (2.0, 17.4) | 15.5 (5.9, 34.4) | 44.6 (18.4, 87.2) | 30.6 (12.4, 61.7) | 28.1 (11.2, 57.0) | 49.8 (20.6, 96.6) |
| | 18–49 years w/ risk conditions[e] | 3.0 (0.9, 7.6) | 6.8 (2.6, 15.1) | 19.5 (8.0, 38.2) | 13.4 (5.4, 27.0) | 12.3 (4.9, 24.9) | 21.8 (9.0, 42.3) |
| | 50–64 years, all[d] | 21.4 (8.4, 44.3) | 38.5 (16.2, 74.1) | 92.4 (40.3, 165.8) | 67.3 (29.1, 123.4) | 57.0 (24.5, 105.3) | 104.3 (45.7, 186.0) |
| | ≥65 years[d,f] | 21.4 (6.9, 50.6) | 72.4 (30.6, 138.7) | 166.7 (73.3, 294.1) | 106.4 (46.0, 195.2) | 95.1 (40.9, 175.3) | 188.0 (82.9, 329.4) |
| | Total, all[g,h] | 50.0 (18.4, 108.3) | 127.7 (54.8, 237.7) | 307.1 (137.3, 522.8) | 206.7 (91.1, 363.8) | 182.3 (80.1, 322.5) | 345.9 (155.2, 584.7) |
| | Total, PCVs recommended[g,i] | 46.1 (17.1, 99.3) | 118.7 (51.0, 220.5) | 281.2 (125.6, 479.6) | 188.9 (83.3, 333.0) | 166.0 (72.9, 294.0) | 317.0 (142.1, 537.0) |

| Condition | Age category | Number of cases in thousands (95% CI)[b] | | | | | |
|---|---|---|---|---|---|---|---|
| | | *PCV15* | *PCV20* | *PCV21* | *PCV24*[c] | *PCV25* | *PCV31* |
| Pneumonia (inpatients)[j] | | | | | | | |
| | 18–49 years, all[d] | 0.2 (0.0, 0.4) | 0.4 (0.1, 0.8) | 1.1 (0.4, 2.0) | 0.7 (0.3, 1.4) | 0.7 (0.3, 1.3) | 1.2 (0.5, 2.3) |
| | 18–49 years w/ risk conditions[e] | 0.1 (0.0, 0.2) | 0.2 (0.1, 0.5) | 0.6 (0.3, 1.2) | 0.4 (0.2, 0.8) | 0.4 (0.2, 0.8) | 0.7 (0.3, 1.3) |
| | 50–64 years, all[d] | 0.8 (0.3, 1.6) | 1.4 (0.6, 2.7) | 3.4 (1.5, 5.9) | 2.5 (1.1, 4.4) | 2.1 (0.9, 3.8) | 3.8 (1.7, 6.6) |
| | ≥65 years[d,f] | 1.1 (0.4, 2.5) | 3.7 (1.6, 6.9) | 8.5 (3.8, 14.7) | 5.4 (2.4, 9.7) | 4.8 (2.1, 8.8) | 9.6 (4.3, 16.4) |
| | Total, all[g,h] | 2.0 (0.7, 4.4) | 5.5 (2.4, 10.2) | 13.0 (5.8, 22.0) | 8.7 (3.8, 15.2) | 7.7 (3.4, 13.5) | 14.7 (6.6, 24.6) |
| | Total, PCVs recommended[g,i] | 2.0 (0.7, 4.3) | 5.3 (2.3, 9.9) | 12.6 (5.6, 21.3) | 8.4 (3.7, 14.7) | 7.4 (3.3, 13.0) | 14.2 (6.4, 23.8) |
| IPD | | | | | | | |
| | 18–49 years, all[d] | 0.6 (0.4, 0.8) | 1.5 (0.9, 1.9) | 2.8 (1.7, 3.4) | 2.2 (1.3, 2.7) | 2.1 (1.3, 2.7) | 3.1 (1.9, 3.8) |
| | 18–49 years w/ risk conditions[e] | 0.4 (0.3, 0.5) | 1.0 (0.6, 1.3) | 1.9 (1.2, 2.4) | 1.5 (0.9, 1.9) | 1.5 (0.9, 1.8) | 2.1 (1.3, 2.6) |
| | 50–64 years, all[d] | 1.0 (0.6, 1.3) | 2.4 (1.4, 2.9) | 4.5 (2.8, 5.6) | 3.3 (2.0, 4.1) | 3.4 (2.1, 4.3) | 7.1 (4.4, 8.8) |
| | ≥65 years[d,f] | 1.5 (0.9, 1.9) | 2.9 (1.8, 3.6) | 6.1 (3.7, 7.6) | 3.6 (2.2, 4.5) | 4.4 (2.7, 5.5) | 6.6 (4.0, 8.2) |
| | Total, all[g,h] | 3.1 (1.9, 3.9) | 6.8 (4.1, 8.4) | 13.5 (8.2, 16.7) | 9.1 (5.5, 11.3) | 10.0 (6.1, 12.4) | 16.8 (10.3, 20.8) |
| | Total, PCVs recommended[g,i] | 2.9 (1.8, 3.6) | 6.3 (3.8, 7.8) | 12.6 (7.7, 15.6) | 8.4 (5.1, 10.4) | 9.4 (5.7, 11.6) | 15.8 (9.7, 19.6) |

Abbreviations: CI – confidence interval; PCV – pneumococcal conjugate vaccine; IPD – invasive pneumococcal disease.

[a] PCV15 includes serotypes 1, 3, 4, 5, 6A/C, 6B, 7F, 9V, 14, 18C, 19F, 19A, 22F, 23F, 33F. PCV20 includes serotypes 1, 3, 4, 5, 6A/C, 6B, 7F, 8, 9V, 10A, 11A, 12F, 14, 15B, 18C, 19F, 19A, 22F, 23F, 33F. PCV21 includes serotypes 3, 6A/C, 7F, 8, 9N, 10A, 11A, 12F, 15A, 15C, 16F, 17F, 19A, 20A, 22F, 23A, 23B, 24F, 31, 33F, 35B. PCV24 includes serotypes 1, 2, 3, 4, 5, 6A/C, 6B, 7F, 8, 9N, 9V, 10A, 11A, 12F, 14, 15B, 17F, 18C, 19F, 19A, 20B, 22F, 23F, 33F. PCV25 includes serotypes 1, 2, 3, 4, 5, 6B, 6A/C, 7F, 8, 9N, 9V, 10A, 12F, 14, 15A, 15B, 16F, 18C, 19F, 19A, 22F, 23F, 24F, 33F, 35B. PCV31 includes serotypes 1, 2, 3, 4, 5, 6A/C, 6B, 7F, 7C, 8, 9N, 9V, 10A, 11A, 12F, 14, 15A, 15B, 16F, 17F, 18C, 19F, 19A, 20B, 22F, 23F, 23A, 23B, 31, 33F, 35B.

[b] Considering US population by age group[22] and assuming disease incidence rates as detailed in **Tables S11–S13**, pneumococcal-attributable disease proportions as detailed in **Table S4**, PCV preventable proportion of pneumococci by condition and age-group as detailed in **Table S15**, PCV effectiveness as detailed in **Table S6,** and complete serotype 15B and 15C cross-protection.

[c] Also includes Pn-MAPS24v.

[d] All adults within age group.

[e] Adults within age group with underlying health conditions placing them at increased risk of pneumococcal disease. Proportions of adults with at risk conditions derived from Grant et al. 2023[11] and detailed in **Table S5**. Vaccination with PCV15 (in addition to 23-valent pneumococcal polysaccharide vaccine [PPSV23]), PCV20, or PCV21 recommended for adults 18–49 years with health conditions placing them at increased risk of pneumococcal disease.[14,15]
[f] Vaccination with PCV15 (plus PPSV23), PCV20, or PCV21 recommended for all adults ≥65 years without contraindications for vaccination.[14,15]
[g] Component values may not sum to total due to rounding.
[h] Total for all adults.
[i] Total for adults 18–49 years with at-risk conditions and all adults ≥50 years.
[j] Non-bacteremic pneumonia. Bacteremic pneumonia classified as IPD.

### Supplemental Material References
